# Genetic risk in extremely early onset type 1 diabetes

**DOI:** 10.64898/2025.12.18.25342362

**Authors:** Amber M. Luckett, Georgia Bonfield, Gareth Hawkes, Harry D. Green, Lauric Ferrat, Clara Domingo-Vila, Timothy I.M. Tree, William A. Hagopian, Bart O. Roep, Michael N. Weedon, Matthew B. Johnson, Stephen S Rich, Richard A. Oram, EXE-T1D consortium

## Abstract

Identifying individuals at risk of early onset type 1 diabetes (diagnosed <2 years) would be highly beneficial in reducing risk of severe diabetic ketoacidosis (DKA) for those with extreme autoimmunity. We aimed to investigate whether genetic variation contributes to heterogeneity in age of type 1 diabetes onset, focusing on those diagnosed <2 years and ages previously defined by histological differences. We carried out association testing on 6773 individuals with type 1 diabetes and tested for heterogeneity in Human Leukocyte Antigen (HLA) variants across stratified age groups (594 diagnosed <2 years, 2241 diagnosed 2-7 years, 3094 diagnosed 7-13 years, 844 diagnosed 13+ years). We used a 67 SNP type 1 diabetes genetic risk score (T1D-GRS) to quantify aggregated genetic risk and assessed its utility in screening for type 1 diabetes <2 years. We observed higher T1D-GRSs as age of onset decreased in type 1 diabetes and found that DR3-DQ2 homozygosity was most strongly associated with <2 years onset (log-OR=4.27). The T1D-GRS showed high discriminative ability for <2 years onset type 1 diabetes onset (AUC=0.94) and correctly identified 88% of type 1 diabetes cases at the 85th population centile. We have shown higher genetic risk for very early onset T1D and suggest T1D-GRSs in newborn screening is likely to be particularly sensitive to those with younger type 1 diabetes onset.

## Introduction

Understanding the mechanisms underlying differences in age of type 1 diabetes is critical for disease prediction, progression and treatment (1). It was previously thought that all diabetes diagnosed in the neonatal period had monogenic aetiology, though recent studies have provided evidence of polygenic type 1 diabetes occurring in early infancy (2). Genetic and histology-based studies have determined differences in genetic variation or islet immune cell profiles within those diagnosed <7 years compared to 13+ years, though the rapid breakdown of self-tolerance in individuals diagnosed <2 years has yet to be understood and may provide evidence of additional heterogeneity in type 1 diabetes (1,3,4).

Genome-wide association studies (GWAS) have highlighted genetic associations contributing to type 1 diabetes age-of-onset (5). The genetic contribution to type 1 diabetes risk can be quantified using a 67 SNP type 1 diabetes genetic risk score (T1D-GRS). The utility of GRSs in screening individuals diagnosed <2 years, who would not be captured by current autoantibody screening timing recommendations, has not been explored (5–10). Identifying individuals at risk of developing diabetes <2 years would be beneficial in signposting individuals to receive additional glucose monitoring or islet autoantibody testing (6,7,10). This early intervention would increase awareness and reduce diabetic ketoacidosis (DKA) events within this vulnerable population (11).

In this study, we performed GWASs to investigate genetic variants which may contribute to age of type 1 diabetes onset, including those diagnosed <2 years who have not been investigated before. We also investigated the contribution of high-risk Human Leukocyte Antigens (HLA) type 1 diabetes variants in the major histocompatibility complex (MHC) within those diagnosed <2 years, 2-7 years, 7-13 years and >13 years to determine the role of these variants which play a key role in autoimmunity. Finally, we assessed T1D-GRS performance in individuals with early-onset type 1 diabetes to determine its utility within screening for these individuals.

## Research Design and Methods

### Research subjects

#### Type 1 Diabetes Genetics Consortium

We performed analysis on a subset of European individuals from the Type 1 Diabetes Genetics Consortium (T1DGC), using a previously defined type 1 diabetes definition (8,12). We analysed Immunochip case-control data from 6480 individuals with type 1 diabetes diagnosed from 6 months-16 years (mean age at onset = 7.8 years) and 9596 controls (Supplementary Table 1), as previously described (8,12).

### The EXtremely Early onset Type 1 Diabetes Study/The Exeter

#### 10,000/Peninsula Research Bank

The EXtremely Early onset Type 1 Diabetes (EXE-T1D) study included 293 individuals with type 1 diabetes, genotyped from whole-blood DNA using the Illumina Global Screening Array; age at onset ranged from birth–62 years (mean age at onset = 2.3 years) (Supplementary Table 1) (13,14). Monogenic diabetes was comprehensively ruled out by sequencing in all individuals diagnosed <9 months, where monogenic diabetes is common (15). We used The Exeter 10,000/Peninsula Research Bank (EXTEND/PRB) cohort as an unselected population cohort (n=11,074), with 10,403 individuals with no known diabetes, using the Illumina Global Screening Array. Further details of EXTEND/PRB recruitment are described by Rodgers et al. (16).

#### UK Biobank

When assessing T1D-GRS population centiles, we used T1D-GRS generated for individuals of European ancestry in UK Biobank (UKB) (n=367,318) (Supplementary Table 1) without type 1 diabetes using published case-control definitions determined from electronic health records (8,9). Our type 1 diabetes definition consisted of several criteria including a clinical type 1 diabetes diagnosis <20 years of age, on insulin within 1 year from the time of diagnosis and no self-report of type 2 diabetes (8,9).

### Quality control and imputation

We had a total of 131,805 genotyped variants from the T1DGC cohort, 693,391 from the EXE-T1D cohort and 725,831 from the EXTEND/PRB cohort. Due to genotyping occurring on the same array, we merged the EXE-T1D and EXTEND/PRB cohort prior to imputation. We applied the HRC preparation program version 4.2.9 (https://www.well.ox.ac.uk/~wrayner/tools/) for strand alignment. We used PLINK version 1.9 (17) and filtered to remove variants with: minor allele frequency (MAF) <1% and genotypes missing in >5%, in violation of Hardy-Weinberg equilibrium (*P*<1×10^-6^), or with allele ambiguity. We removed samples with: >5% missing genotypes; sex mismatch between genetically defined and reported phenotypes; and removed samples based on pairwise identical-by-descent (IBD) estimates generated with KING version 2.2.4. We performed a Principal Component Analysis (PCA) using GCTA version 1.94.1, using the 1000 Genomes Project (1KGP) reference panel for PC projection (18).

We used the TopMED Imputation Server R3 panel for genome-wide imputation, imputing 442,877,347 variants for T1DGC and 421,250,241 variants for EXE-T1D and EXTEND/PRB (19,20). We imputed a total of 22,733 HLA variants for all cohorts using the Michigan imputation server multi-ethnic HLA reference panel (19). We performed post-imputation filtering to keep variants with: MAF >1%; genotype imputation quality (INFO score) >0.4; and imputation R-squared accuracy R^2^ >0.3 for genome-wide imputed variants, and R^2^ <0.5 for HLA imputed variants.

### Association testing

For all association testing we used REGENIE version 3.2.6 (21). In the T1DGC cohort, we first performed case-control association testing stratified by age at diagnosis (<7 years, n=2560 vs. >13 years, n=836) to identify potential genetic differences in age groups suggested by prior studies. We then performed association testing using age at onset as a continuous trait to assess genetic variation impacting timing of onset rather than overall disease risk. Phenotypes were rank-based inverse normalized (--rint) in weeks to standardise distributions and enable meta-analysis across cohorts.

For the EXE-T1D cohort (n=293), only continuous age-at-onset association testing was performed, as the study design lacked appropriate controls for case-control analysis. Inclusion of EXE-T1D increased power for identifying genetic variants affecting age at onset, particularly in those diagnosed <2 years. To combine results from the continuous age-at-onset GWAS, we conducted an inverse-variance weighted fixed-effects meta-analysis using METAL (22). Effect sizes and standard errors from each cohort were combined using the “SCHEME STDERR” option, weighting by the inverse of the squared standard error. Summary statistics were filtered to include variants with INFO>0.8 and MAF>0.01 in the T1DGC GWAS, and MAF>0.05 in the EXE-T1D cohort to account for its smaller sample size. We identified lead variants using PLINK version 1.9 linkage disequilibrium (LD)-based clumping (adjusting the index SNP significance threshold to 5×10^-8^).

We removed HLA variants and calculated the genomic inflation (λgc) for the association tests (<7 years type 1 diabetes GWAS λgc=0.99, >13 years type 1 diabetes GWAS λgc=1.14, T1DGC continuous GWAS λgc=0.98, EXE-T1D continuous GWAS λgc=0.99, meta-analysed GWAS λgc=0.99).

### Heritability and genetic correlation

We used GCTA version 1.94.1 to perform a bivariate Genome-based Restricted Maximum Likelihood (GREML) analysis to estimate the variance, heritability and genetic correlation of type 1 diabetes diagnosed <7 years and diagnosed >13 years in the T1DGC cohort. We used an estimated prevalence of 0.4% for both analyses based on published estimates (4).

### Heterogeneity testing

We tested heterogeneity using a Firth regression heterogeneity test (PLINK version 1.9) including PCs 1-5 and sex as covariates. We tested the difference in effect size in type 1 diabetes diagnosed <7 years compared to >13 years within genome-wide significant loci (excluding MHC region due to high LD between HLA loci) and in 67 SNPs present in the T1D-GRS2x model (to determine if known type 1 diabetes loci have significant heterogeneity despite not reaching genome-wide significance).

### HLA loci contribution to type 1 diabetes onset

From the imputed HLA haplotypes, we determined carrier status for Class I and Class II HLA variants found to be heterogeneous in our case-control association tests and also found to be associated with age of type 1 diabetes onset from previously published studies (4). To determine the role of HLA within early onset type 1 diabetes we first performed logistic regressions comparing each HLA allele across age groups using the youngest group (<2 years) as the reference. Alleles showing significant differences were then included in a multinomial regression comparing all age-of-onset groups to controls, adjusting for other significant HLA alleles. Class II HLA haplotypes were collapsed into a category (“DRB1 genotype”) to avoid collinearity between homozygotes and single copy carriers (with “X” conveying another Class II HLA allele not present in any other group) (Supplementary Table 2). All regression analyses were adjusted for the first 5 principal components, sex and genotype array used. Multiple testing was controlled using the Benjamini-Hochberg false discovery rate. We also performed a sensitivity analysis, adjusting for interactions between significant HLA alleles, to ensure associations were not driven by HLA interactions.

### Genetic risk scores

We used the python PRSedm package (https://github.com/sethsh7/PRSedm) (23) to generate polygenic risk scores for all individuals in our cohorts (9). EXTEND/PRB participants were included as a control group for the EXE-T1D cohort, as both were genotyped on the same array and imputed simultaneously. We used a logistic regression model to test for associations between GRS (predictor) and type 1 diabetes status (outcome) using sex,the first 5 PCs, and genotype array as covariates.

We generated partitioned type 1 diabetes polygenic risk scores using the 67-SNP type 1 diabetes GRS2x model (9). We assessed differences in T1D-GRS by age of onset, using established age groups and individuals diagnosed <2 years.

### Ethics

The Extremely Early-Onset Type 1 Diabetes (EXE-T1D) study has ethical approval from Derby Research Ethics Committee, Derby, U.K. (IRAS project ID 228082). The EXTEND/PRB study has ethical approval from Southwest-Cornwall & Plymouth NHS Research Ethics Committee, Bristol, U.K. (reference 14/SW/1089; 5-year extension following initial approval: reference 09/H0106/75).

## Data and Resource Availability

GRS2x along with relevant diabetes polygenic scores are available through the Polygenic Risk Score Extension for Diabetes Mellitus (PRSedm) package and hosted on Zenodo (https://zenodo.org/records/17903390). This research utilised data from the UK Biobank resource carried out under UK Biobank application number 103356. Data from the T1DGC are accessible from public repositories: NIDDK Central Repository (https://repository.niddk.nih.gov/home); the database of Genotypes and Phenotypes (dbGaP, https://dbgap.ncbi.nlm.nih.gov/aa/wga.cgi?page=login, accession Nos. phs000180, phs000910, phs000911, phs001222, phs001426, and phs002468); the European Genome-Phenome Archive (EGA, https://ega-archive.org/); and the T1D Knowledge Portal (https://t1d.hugeamp.org/).

## Results

### Genome-wide association study of case-control groups

We stratified the 6480 type 1 diabetes cases based on a type 1 diabetes diagnosis <7 years (n= 2560) and >13 years (n= 836) and compared against 9599 controls from T1DGC.

We identified 4 genome-wide significant loci (*P*<5 × 10□□) in the <7 years type 1 diabetes onset group and 2 loci in the >13 years onset group. For individuals diagnosed <7 years, we observed significant associations at MHC (lead SNP: rs3873448, *P*=2.23×10^-242^), *INS* (lead SNP: rs3842753, *P*=1.87×10^-59^), *PTPN22* (lead SNP: rs2476601, *P*=2.14×10^-30^), and *LINC01882* (lead SNP: rs67878610, *P*=1.64×10^-11^), all of which have been previously reported in association with type 1 diabetes (24). For individuals with a diagnosis >13 years, we detected significant associations at the MHC (lead SNP: rs1794269, *P*=1.65×10^-89^ and *INS* (lead SNP: rs689, *P*=1.71×10^-19^) loci (Supplementary Fig. 1).

### Heterogeneity in variant effects in <7 years type 1 diabetes diagnosis and >13 years type 1 diabetes diagnosis

We next tested heterogeneity between variant effects in <7 years and >13 years type 1 diabetes onset. Of the 4 genome-wide significant loci we found, no loci remained significant (*P*>0.05) (Supplementary Fig. 2). We then tested for heterogeneity in T1D-GRS variant effect sizes across the <7 year and >13 year onset groups. Among 67 SNPs (including tagged HLA haplotypes with established linkage from the type 1 diabetes GRS2x model), 15 loci demonstrated evidence of heterogeneity at nominal significance (*P*< 0.05) (Supplementary Fig. 3 and Supplementary Table 3). No variants remained significant at false discovery rate (FDR) <0.1, though HLA variants significant at FDR<0.2 were included in downstream analyses.

### Genetic variance, heritability and correlation

We next estimated the heritability (on the liability scale) explained by SNPs in the <7 year and >13 year type 1 diabetes onset groups. GCTA-GREML analysis further highlighted the greater genetic component within the <7 years onset group. The heritability was greater in <7 years onset group (h^2^=0.42, SE=0.01) compared to the >13 years group (h^2^=0.24, SE=0.06) (*P*=3.1×10^-3^), with the estimated genetic correlation (r_g_) between the two groups at 0.71 (SE=0.12) (Supplementary Fig. 4), suggestive of a difference in genetic contribution between the traits.

### Association testing using age-at-onset as continuous phenotype

The association testing using type 1 diabetes onset as a continuous phenotype yielded no significant (*P*<5×10^-8^) associations. From the EXE-T1D cohort, 9 loci had suggestive evidence of association (*P*< 1×10□□); *RN7SL807P* (rs1517628, *P*=1.7×10^-6^), *FAM47E* (rs10025494, *P*=7.7×10-6), *EDIL3* (rs2221761, *P*=2.1×10^-6^), *ENSG00000300630* (rs9406047, *P*=3.0×10^-6^), *NTM* (rs139416102, *P*=4.7×10^-6^), *STON2* (rs61978878, *P*=4.9×10^-6^), *RORA* (rs12592002, *P*=8.2×10^-6^), DYNLRB2-AS1 (rs1878821, *P*=6.5×10^-6^), and *ENSG00000304175* (rs79505903, *P*=3.3×10^-6^) (Supplementary Fig. 5A). In the T1DGC cohort, one loci reached this threshold; *MIR4300HG* (rs12792418 *P*=1.6×10^-6^) (Supplementary Fig. 5B). After meta-analysis, 8 signals had suggestive evidence of association (Supplementary Fig. 6). Notably, these signals were only present in the EXE-T1D cohort association testing due to INFO and MAF filtering removing them from the T1DGC cohort; therefore, the meta-analysis reflected evidence solely from the EXE-T1D cohort.

### HLA variant contribution to age of type 1 diabetes onset

Of the 19 HLA loci showing potential heterogeneity in type 1 diabetes age of onset (based on our heterogeneity tests and prior studies), seven loci were nominally significantly associated with early-onset type 1 diabetes in initial logistic regression analyses (Supplementary Table 4).

We next assessed the effects of these HLA loci on age at onset using multinomial regression, with controls as the reference group. All DRB1 genotypes stratified by age at onset were significantly different from the control group at *Padj*<2.2×10^-16^ (Fig. 1). The *DR3-DQ2*/*DR4-DQ8* and *DR3-DQ2*/*DR3-DQ2* loci were consistently associated with the youngest onset group (<2 years), showing the highest log-odds ratios (log-OR) (*DR3-DQ2*/*DR4-DQ8* log-OR = 5.05; *DR3-DQ2*/*DR3-DQ2* log-OR =4.27) (Fig. 1). The *DR4-DQ8*/X and *DR3-DQ2*/X genotypes also conferred elevated risk in younger age groups, though with log-OR (*DR4-DQ8*/X log-OR = 2.33; *DR3-DQ2*/X log-OR = 2.07 (Fig. 1).

**Fig. 1.**
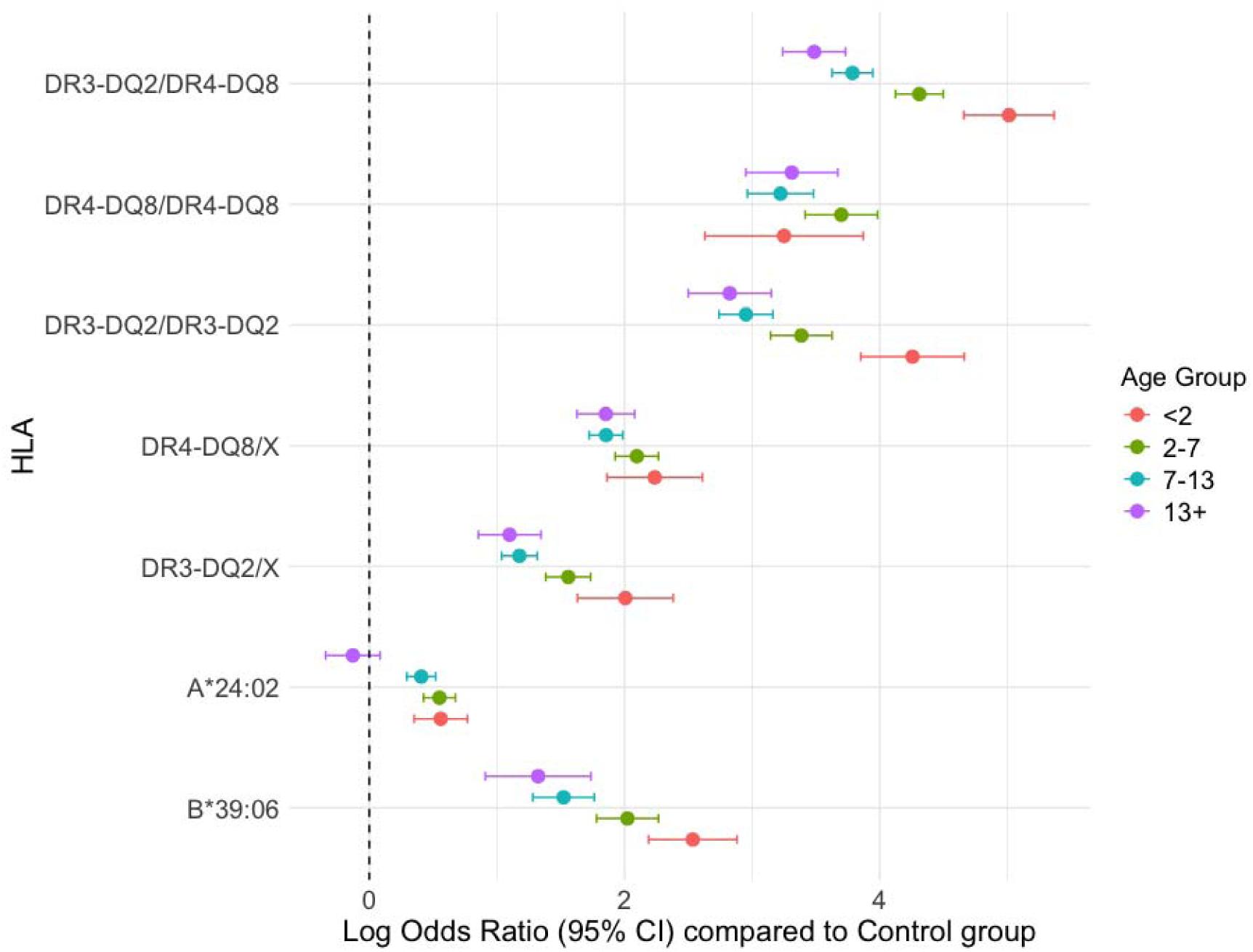
Log-Odds ratios of *HLA* alleles and haplotypes at different type 1 diabetes onsets. Log Odds Ratios (log-OR) and 95% Confidence Intervals (CI) for selected HLA Class II diplotypes (DR3-DQ2/DR4-DQ8, DR4-DQ8/DR4-DQ8, DR3-DQ2/DR3-DQ2), major haplotypes (DR4-DQ8/X, DR3-DQ2/X where “X” represents a Class II HLA allele independent of the DR3-DQ2 and DR4-DQ8 haplotype), and Class I alleles (A*24:02, B*39:06) compared to the control group (log-OR = 0). Alleles were assessed using a multinomial regression model adjusted for the first five Principal Components (PCs), sex, and genotyping array. The results are stratified by age of T1D onset: <2 years (red), 2-7 years (green), 7-13 years (blue), and 13+ years (purple).

Among class I alleles, *B*39:06* and *A*24:02* were associated with earlier onset (log-OR = 2.53, *Padj*<2.2×10^-16^; log-OR = 0.49, *Padj*=5.1×10^-6^), while in the oldest onset group, A*24:02 carriers showed no significant difference compared with controls (*P*=0.17) (Fig. 1). All associations that were significantly different from controls remained significant at FDR<0.1.

A sensitivity analysis including interaction terms between all significant HLA loci was performed to determine the influence of interaction effects on HLA contribution to type 1 diabetes onset. Across 44 tested pairwise interactions, 35 showed no effect on HLA loci additive effect (*Padj*> 0.05), indicating largely independent effects of these loci (Supplementary Table 5). Loci interactions with significant associations include *A24:02* × *DR3-DQ2*/X in the 7–13 years onset group (*Padj*=6.7×10^-3^) and several *B39:06* × DRB1 genotype combinations (e.g., *B39:06* × *DR3-DQ2*/X in 2–7 [*Padj*=0.049], and 7–13 [*Padj*=0.036,] and 13+ age groups [*Padj*=0.022]; *B39:06* × *DR3-DQ2*/*DR4-DQ8* in <2 [*Padj*=0.012], 2–7 [*Padj*=2.89×10□□], 7–13 [*Padj*=8.8×10^-4^], and 13+ groups [*Padj*=0.016]). Notably, *A*24:02* additive effects were generally non-significant in this sensitivity analysis, suggesting that its contribution to type 1 diabetes onset may be influenced by other HLA alleles. Some extreme OR estimates (e.g., *B*39:06* × *DR3-DQ2*/*DR3-DQ2* and *B*39:06* × *DR4-DQ8*/*DR4-DQ8*) are likely driven by sparse data in certain genotype-age strata (n= 15 and n=17, respectively), though biological interactions have been previously detailed (25).

### Type 1 Diabetes Genetic Risk Scores

We found that individuals diagnosed <2 years had the highest T1D-GRS (mean=15.1, 95% CI 15.0-15.3) compared to diagnosis at: 2-7 years (mean=14.7, 95% CI 14.6-14.8; *Padj*= 4.14×10^-6^); 7-13 years (mean=14.4, 95% CI 14.3-14.5; *Padj*=3.27×10^-15^); 13+ years (mean=14.1, 95% CI 14.0-14.2; *Padj*= 1.46×10^-19^); and compared to the control group (mean= 10.5, 95% CI 10.5-10.6; *Padj*= 4.40×10^-16^) (Fig. 2 and Supplementary Fig. 7). The higher T1D-GRS in the <2 years group was driven by the Class II HLA variant interaction score and the Class I HLA variant interaction score (Supplementary Fig. 8)

**Fig. 2.**
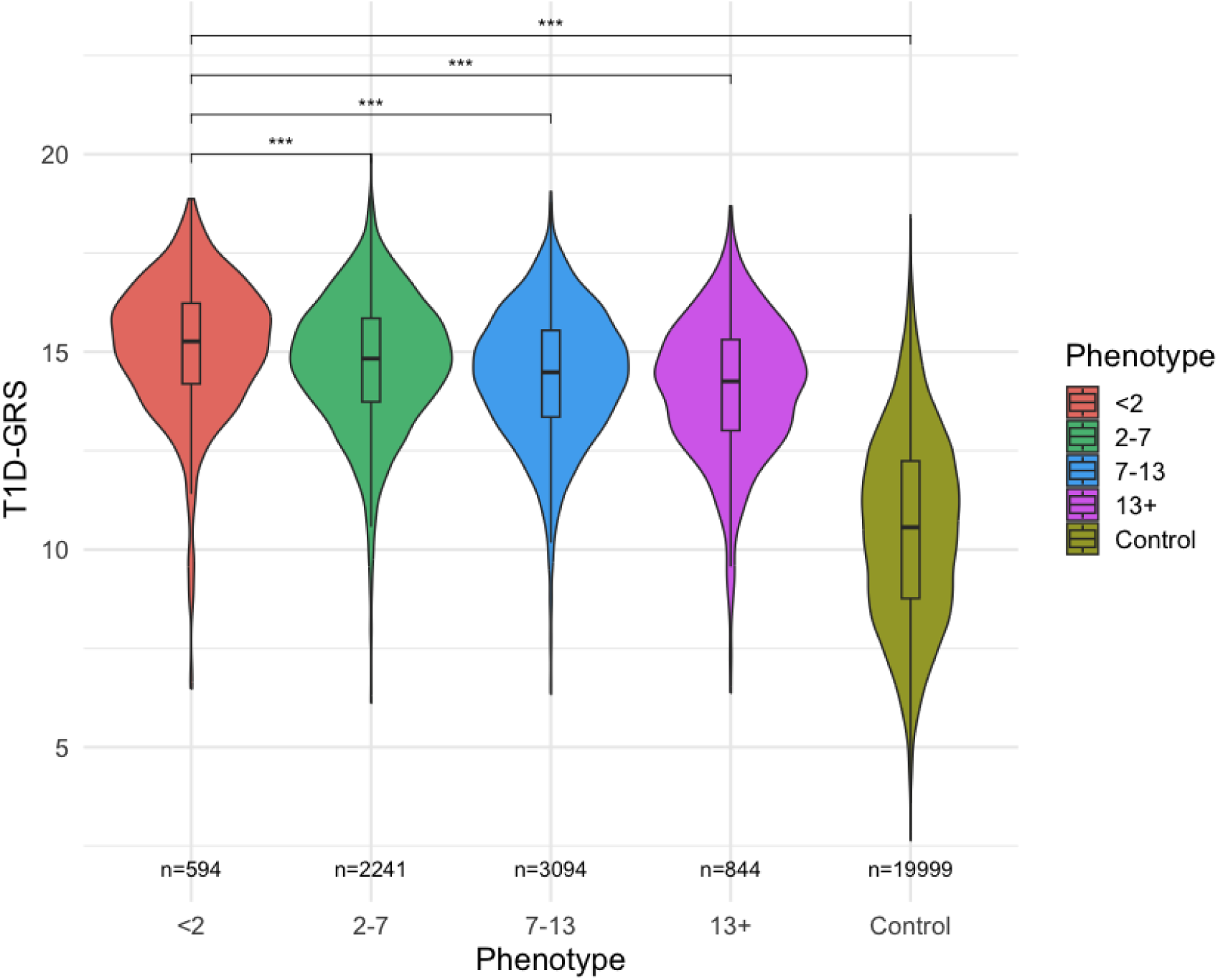
T1D-GRS Distribution across different diagnosis groupings in T1DGC and EXE-T1D/EXTEND Cohorts. Violin plots show the distribution of the 67-SNP T1D-GRS for type 1 diabetes cases stratified by age of diagnosis compared to controls. The box plot within each violin shows the median, interquartile range (IQR), and 95% confidence intervals. All *P*-values derived from logistic regression adjusting for first five Principal Components (PCs),sex, and array used.

### T1D-GRS discriminative performance and application in screening

The T1D-GRS demonstrated strong discriminative ability for identifying individuals with type 1 diabetes onset <2 years, largely driven by Class II HLA variants (Supplementary Figure 9). Receiver Operating Characteristic (ROC) analysis comparing type 1 diabetes diagnosed <2 years to controls yielded an Area Under the Curve (AUC) of 0.94 (95% CI 0.93–0.95), indicating high predictive accuracy. When comparing early-onset (<2 years) to later childhood-onset (>2 years) type 1 diabetes, the GRS2 showed moderate discrimination (AUC=0.62 [95% CI 0.60–0.65]), consistent with a gradient of genetic risk by age at onset (Fig. 3).

**Fig. 3.**
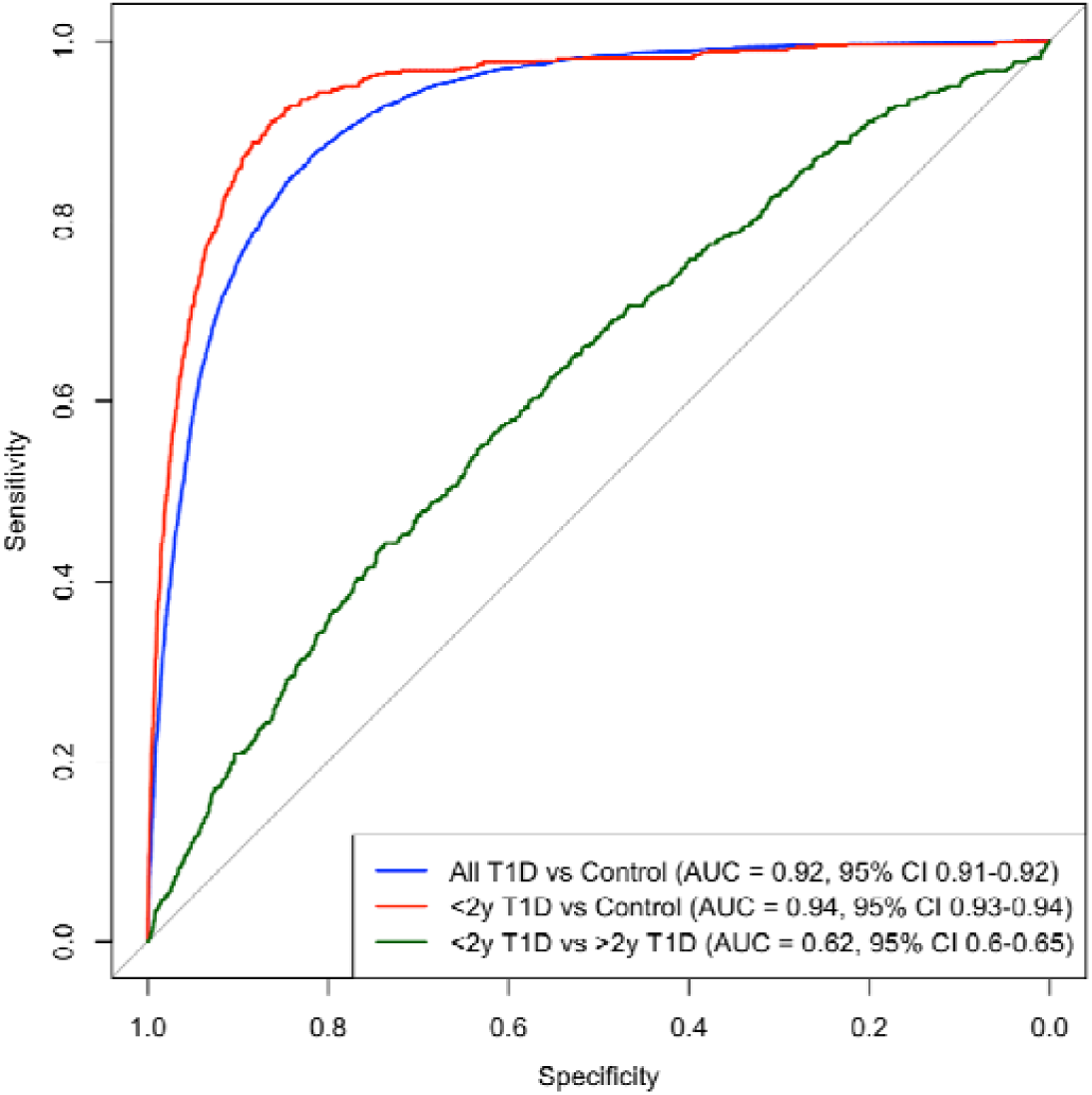
Receiver Operating Characteristic (ROC) Curves for T1D-GRS Discriminative Performance. ROC curves of the 67-SNP T1D-GRS discriminative ability between type 1 diabetes (T1D) phenotypes: all T1D cases vs. controls (AUC=0.92, 95% CI 0.91-0.92) (blue), extremely early-onset T1D (<2 years) vs. controls (red) (AUC=0.94, 95% CI 0.93-0.94), <2 years T1D vs. >2 years T1D onset (green) (AUC=0.62, 95% CI 0.6-0.65).

Analysis of T1D-GRS performance across population centile thresholds demonstrated strong discrimination for identifying type 1 diabetes, particularly for individuals diagnosed <2 years. Of the thresholds tested, the optimal balance between sensitivity and specificity was observed at the 85th population centile (T1D-GRS = 12.599; Youden index = 0.68), identifying 88% of all future type 1 diabetes cases with a type 1 diabetes risk of 1.3% (Table 1). In our cohort of children diagnosed <2 years, 94.3% had a T1D-GRS above this cutoff (Supplementary Table 6) with consistent results across both cohorts Supplementary Table 7, Supplementary Table 8) (26).

**Table 1.** Discriminative Performance of T1D-GRS Across Population Centile Thresholds in all type 1 diabetes (T1D). Risk of type 1 diabetes calculated using a 0.3% population prevalence of type 1 diabetes (8). Population centile calculated from UK Biobank European population.

| Population Centile | T1D Centile | T1D-GRS | Sensitivity (%) | Specificity (%) | 1-Specificity (%) | Youden index | T1D risk (%) |
| --- | --- | --- | --- | --- | --- | --- | --- |
| 50 | 2.0 | 10.159 | 98.7 | 44.1 | 55.9 | 0.427 | 0.53 |
| 75 | 6.9 | 11.815 | 94.3 | 69.0 | 31.0 | 0.634 | 0.91 |
| 80 | 9.1 | 12.185 | 92.0 | 74.3 | 25.7 | 0.663 | 1.07 |
| 85 | 13.0 | 12.599 | 88.0 | 79.9 | 20.1 | 0.680 | 1.30 |
| 90 | 20.6 | 13.115 | 81.2 | 86.0 | 14.0 | 0.672 | 1.72 |
| 95 | 35.0 | 13.882 | 67.6 | 92.3 | 7.7 | 0.600 | 2.57 |
| 99 | 66.9 | 15.303 | 33.6 | 98.0 | 2.0 | 0.316 | 4.81 |
| 100 | 100 | 19.103 | 0 | 100 | 0 | 0 | NA |

## Discussion

To our knowledge, we have performed the largest exploratory genetic analysis focusing on individuals diagnosed with type 1 diabetes at <2 years. We have highlighted the strong genetic risk associated with this phenotype by showing that those with the youngest onset carry the highest aggregate type 1 diabetes genetic risk. We have also demonstrated how we can utilise GRSs to screen for this rare but clinically important phenotype.

The increased genetic contribution to earlier onset Type 1 Diabetes was evident from our heritability analyses as well as the genetic risk score calculated. Previous studies have detailed the inverse relationship between age of onset and type 1 diabetes genetic risk (2,8). We further support this finding, with the <2 year onset group having higher genetic risk than other groups (*P*<0.05). We found a significantly increased heritability in the <7 years age group (h^2^=0.42) when compared to the >13 years group (h^2^=0.24). This heritability estimate for the early-onset group is higher than the previously published estimate for the same age group (h^2^=0.37), while the estimate for the older age group remained consistent (h^2^=0.23), likely due to heritability estimates being ascertained from imputed variants rather than using chip-based estimates (4). By quantifying a genetic correlation between these groups, we were able to further highlight how age stratified type 1 diabetes based on histologically defined groups is less genetically similar (r_g_=0.71) to estimates between sex-stratified type 1 diabetes (r_g_=0.88) (though more similar than high-risk HLA stratified type 1 diabetes [r_g_=0.68]) (27).

Although the genetic contribution is clear, we did not identify novel independent loci specifically associated with type 1 diabetes onset in our association testing, suggesting that the genetic differences are more likely due to the aggregation of multiple, common risk alleles (as seen in the increased mean T1D-GRS in the <2 years groups) rather than the effect of single variants. Our HLA heterogeneity analyses revealed further differences in genetic contribution to type 1 diabetes onset. The *DR3-DQ2*/*DR4-DQ8* diplotype is well-established as conveying the highest overall type 1 diabetes risk (OR=37.80) (8), and had the largest contribution to type 1 diabetes <2 years (log-OR= 5.05). We found that within homozygous combinations, DR3-DQ2 homozygosity was more strongly associated with type 1 diabetes onset at <2 years (log-OR= 4.27) compared to other homozygous HLA loci. Although *DR4-DQ8* homozygosity is more associated with childhood-onset type 1 diabetes (28) (as we found in ages 2-13+ years), our study suggests that *DR3-DQ2* homozygosity predominates within the <2 years-onset group. Further biological and immunological studies could help to determine the impact of the *DR3-DQ2* homozygous haplotype on type 1 diabetes pathogenesis and its role in the rapid progression of type 1 diabetes.

Clinically, T1D-GRS demonstrated high discriminative ability (AUC = 0.94) for identifying infants with early-onset T1D. Implementation of GRS-based newborn screening could capture high-risk infants who would benefit from close monitoring and islet autoantibody testing, reducing the incidence of diabetic ketoacidosis at diagnosis (29). Using a T1D-GRS threshold at the 85^th^ population centile yielded the optimum Youden index (0.68), correctly identifying 88% of eventual type 1 diabetes and ∼94% of type 1 diabetes diagnosed <2 years in our cohorts. Further work is required for validating cost-effectiveness and feasibility in population settings, assessing ethical implications, and ensuring equitable performance across ancestries (29). Our study is limited by our cohorts mainly consisting of individuals with White European ancestry and therefore results may not be translatable across ancestries. Additional studies utilising cohorts with varying ancestries would be highly beneficial in determining T1D-GRS utility combined with islet autoantibody testing in screening prior to clinical translation.

## Conclusions

In summary, individuals with an earlier onset of type 1 diabetes have a higher aggregated genetic risk. Additionally, we found evidence that *DR3-DQ2* homozygosity conveys the greatest risk in type 1 diabetes diagnosed <2 years, compared to the role of *DR4-DQ8* in later childhood onset, suggesting key autoimmune loci contribute to differences in type 1 diabetes onset. The high discriminative performance of T1D-GRS in <2 years type 1 diabetes onset highlights how implementing GRSs into newborn screening is likely to be highly useful for highlighting a subset of individuals who would benefit from follow-up; however further work is required before this can be implemented in clinical practice.

## Supporting information

Authors listed in Supplementary Material

## Data Availability

EXE-T1D and EXTEND/PRB data produced in the present study are available upon reasonable request to the authors. T1DGC data is publically available online via dbGaP, NIDKK Central repository, the European Genome-Phenome Archive, and the T1D Knowledge Portal.

https://repository.niddk.nih.gov/home

https://dbgap.ncbi.nlm.nih.gov/aa/wga.cgi?page=login

https://ega-archive.org/

https://t1d.hugeamp.org/

## Acknowledgments

The authors thank the patients, their relatives, and their physicians for participating in this study. This study was supported by the University of Exeter, the National Institute for Health and Care Research (NIHR) Exeter Biomedical Research Centre, the NIHR Exeter Clinical Research Facility (a partnership between the University of Exeter and Royal Devon University Healthcare NHS Foundation Trust), the NIHR Oxford Biomedical Research Centre, and the NIHR Birmingham Biomedical Research Centre. The views expressed are those of the authors and not necessarily those of the NIHR or the Department of Health and Social Care.

The views expressed are those of the authors and not necessarily those of the National Institute for Health and Care Research or the Department of Health and Social Care.

## Funding

This study was funded by The Leona M. and Harry B. Helmsley Charitable Trust (grants 2016PG-T1D049, 2018PG-T1D049, 2103–05059, and G-2404-06858). M.B.J. is a Diabetes UK and Breakthrough T1D (formerly Juvenile Diabetes Research Foundation) RD Lawrence Fellow (23/0006516). R.A.O. has received support from NIH R01 DK121843-01 and is supported by a JDRF strategic research agreement (3-SRA-2019-827-S-B). R.A.O. and M.N.W. had a U.K. Medical Research Council Confidence in Concept grant to develop a type 1 diabetes GRS biochip with Randox and have ongoing research funding from Randox R&D. W.A.H. is supported by U.S. National Institutes of Health grant 5U01DK128847 and by an unrestricted research grant from Sanofi U.S. A.M.L. is funded by a PhD studentship from Randox Laboratories Ltd. The T1DGC was a collaborative clinical study sponsored by the NIDDK, National Institute of Allergy and Infectious Diseases (NIAID), National Human Genome Research Institute (NHGRI), Eunice Kennedy Shriver National Institute of Child Health and Human Development (NICHD), and the JDRF and supported by U01 DK-062418.

## Duality of Interest

R.A.O. has served as a consultant for Sanofi Pharmaceuticals, ProventionBio, and Janssen Pharmaceuticals. R.A.O. and M.N.W. had a U.K. Medical Research Council confidence-in-concept award to develop a type 1 diabetes Genetic Risk Score biochip with Randox R & D and have ongoing research funding from Randox Laboratories Ltd. A.M.L. is funded by a PhD studentship from Randox Laboratories Ltd. No other potential conflicts of interest relevant to this article were reported.

## Author Contributions

A.M.L. wrote the manuscript and researched the data. G.H., H.D.G., L. F., C.D-V., T.I.M.T., B.O.R., and W.A.H contributed to the methods. B.O.R., M.N.W., and R.A.O. reviewed/edited the manuscript and contributed to the discussion. M.B.J., and G.B contributed to data handling. S.S.R. contributed resources and reviewed the manuscript. R.A.O. is the guarantor of this work and, as such, had full access to all the data in the study and takes responsibility for the integrity of the data and the accuracy of the data analysis.

## Notes

### Author Declarations

The Extremely Early Onset Type 1 Diabetes (EXE-T1D) study has ethical approval from Derby Research Ethics Committee, Derby, U.K. (IRAS project ID 228082). The EXTEND/PRB study has ethical approval from Southwest Cornwall & Plymouth NHS Research Ethics Committee, Bristol, U.K. (reference 14/SW/1089; 5-year extension following initial approval: reference 09/H0106/75). All necessary approvals were obtained from the Type 1 Diabetes Genetics Consortium under a data use agreement with the University of Virginia. This research utilised data from the UK Biobank resource carried out under UK Biobank application number 103356.

