## Supplementary material for "Genetic risk in extremely early onset type 1 diabetes": Authors listed in Supplementary Material

EXE-T1D Consortium members:

Department of Clinical & Biomedical Sciences, University of Exeter Medical School, Exeter, UK

Rebecca A Dobbs, Andrew T Hattersley, Michelle Hudson, Timothy J McDonald, Noel G Morgan, Kathryn Murrall, Sarah J Richardson, Suraj Ramchand, Dill Patel

Department of Immunobiology, School of Immunology & Microbial Sciences (SIMS), King's College London, UK

Paul Blair,

Center for Interventional Immunology, Benaroya Research Institute, Seattle, WA

Bradford Dimos, Cate Speake, Megan E Smithmyer

Translational Health Sciences, Bristol Medical School, University of Bristol, Southmead Hospital, Bristol, UK

Kathleen M Gillespie

Diabetes and Inflammation Laboratory, Centre for Human Genetics, Nuffield Department of Medicine, Bukhman Centre for Research Excellence in Type 1 Diabetes, Oxford NIHR Biomedical Research Centre, University of Oxford, Oxford, UK

Rachel E J Besser

Department of Paediatric Endocrinology and Diabetes, Oxford University Hospitals NHS Foundation Trust, Oxford, UK.

Rachel E J Besser

Department of Paediatric Endocrinology and Diabetes, Birmingham Women's and Children's NHS Foundation Trust, Birmingham, UK

Renuka P Dias

Mendy Korer - Participant Parent Representative
