## Supplementary Table 1 for "Genetic risk in extremely early onset type 1 diabetes"

**Supplementary Table 1.** Cohort characteristics.

| Cohort | Total (n) | Total T1D (n) | Total Controls (n) | Mean T1D onset (years) (95% CI) | <2y T1D onset (n) | 2y-7y T1D onset (n) | 7y-13y T1D onset (n) | 13y+ T1D onset (n) | % Females T1D | % Females Controls | % European ancestry |
|---|---|---|---|---|---|---|---|---|---|---|---|
| **EXET1D/ EXTEND/ PRB** | 10,696 | 293 | 10,403 | 2.3 (1.7–2.9) | 242 | 33 | 10 | 8 | 44 | 50.5 | 97 |
| **T1DGC** | 16,076 | 6480 | 9596 | 7.8 (7.7–7.8) | 352 | 2208 | 3084 | 836 | 49.3 | 51.5 | 100 |
| **UKB** | 367,318 | NA | 367,318 | NA | NA | NA | NA | NA | NA | 54.5 | 100 |
