## Supplementary Table 2 for "Genetic risk in extremely early onset type 1 diabetes"

**Supplementary Table 2.** Classification of HLA haplotypes and associated abbreviation.

| **DR-DQ haplotype** | **Abbreviation** |
|---|---|
| DRB1*07:01-DQA1*02-DQB1*02:02 | DQ2.2 |
| DRB1*03:01-DQA1*05-DQB1*02:01 | DR3-DQ2 |
| DRB1*08:01-DQA1*04-DQB1*04:02 | DQ4.2 |
| DRB1*01:0X-DQA1*01-DQB1*05:01 | DQ5.1 |
| DRB1*16:0X-DQA1*01-DQB1*05:02 | DQ5.2 |
| DRB1*14:01-DQA1*01-DQB1*05:03 | DQ5.3 |
| DRB1*15:02-DQA1*01-DQB1*06:01 | DQ6.1 |
| DRB1*15:01-DQA1*01-DQB1*06:02 | DQ6.2 |
| DRB1*13:01-DQA1*01-DQB1*06:03 | DQ6.3 |
| DRB1*13:02-DQA1*01-DQB1*06:04 | DQ6.4 |
| DRB1*13:02-DQA1*01-DQB1*06:09 | DQ6.9 |
| DRB1*11:0X-DQA1*05-DQB1*03:01 | DQ7.5 |
| DRB1*08:03-DQA1*06-DQB1*03:01 | DQ7.6 |
| DRB1*04:0X-DQA1*03-DQB1*03:02 | DR4-DQ8 |
| DRB1*07:01-DQA1*02-DQB1*03:03 | DQ9.2 |
| DRB1*09:01-DQA1*03-DQB1*03:03 | DQ9.3 |
| DRB1*04:0X-DQA1*03-DQB1*03:01 | DQ7.3 |
