## Supplementary Table 3 for "Genetic risk in extremely early onset type 1 diabetes"

**Supplementary Table 3.** Effect sizes of HLA variants from <7 year T1D GWAS and >13 year T1D GWAS and P-value from heterogeneity test.

| Locus | <7 year effect size (ß) | >13 year effect size (ß) | ***P*-value** |
|---|---|---|---|
| *DQ6.3* | -2.42 | -1.05 | 1.50E-02 |
| *A*29:02* | -0.38 | -0.04 | 4.30E-02 |
| *B*18:01* | -0.59 | -0.02 | 4.00E-02 |
| *B*45:01* | -0.77 | 0.15 | 1.80E-02 |
| *DQ6.9* | -1.67 | -0.8 | 3.70E-02 |
| *DPB1*15:01* | 0.51 | 0.32 | 3.90E-03 |
| *RBM17* | -0.64 | -0.49 | 4.60E-02 |
| *DQ4.2* | 0.31 | 0.03 | 4.50E-02 |
| *B*39:06* | 1.99 | 0.89 | 6.50E-03 |
| *BNTNL2-DRA1* | 0.56 | 0.53 | 2.10E-02 |
| *B*57:01* | 0.63 | 0.48 | 4.70E-02 |
| *DRA1-DRB1* | 1.75 | 1.11 | 5.70E-03 |
| *DQ9.3* | 0.18 | 0.16 | 2.60E-02 |
| *ITGB7* | 0.22 | 0.41 | 4.70E-02 |
| *A*02:05* | 0.55 | 0.63 | 9.30E-03 |
