## Supplementary Table 4 for "Genetic risk in extremely early onset type 1 diabetes"

**Supplementary Table 4.** Regression output comparing HLA contribution to <2 years type 1 diabetes onset.

| Allele | Phenotype | OR | SE | Z | *P*-value |
|---|---|---|---|---|---|
| HLA A*24:02 | Control | 0.903 | 0.017 | -6.122 | 9.25E-10 |
| HLA A*24:02 | 2-7 | 0.993 | 0.018 | -0.381 | 0.703 |
| HLA A*24:02 | 7-13 | 0.957 | 0.018 | -2.469 | 0.0136 |
| HLA A*24:02 | 13+ | 0.883 | 0.021 | -5.844 | 5.1E-09 |
| HLA A*24:02 | Control | 0.903 | 0.017 | -6.122 | 9.25E-10 |
| HLA A*24:02 | 2-7 | 0.993 | 0.018 | -0.381 | 0.703 |
| HLA A*24:02 | 7-13 | 0.957 | 0.018 | -2.469 | 0.0136 |
| HLA A*24:02 | 13+ | 0.883 | 0.021 | -5.844 | 5.1E-09 |
| DR3 | Control | 0.950 | 0.018 | -2.815 | 0.00487 |
| DR3 | 2-7 | 0.970 | 0.020 | -1.548 | 0.122 |
| DR3 | 7-13 | 0.962 | 0.019 | -2.006 | 0.0448 |
| DR3 | 13+ | 0.967 | 0.023 | -1.474 | 0.14 |
| DR4 | Control | 0.966 | 0.017 | -2.044 | 0.041 |
| DR4 | 2-7 | 1.092 | 0.019 | 4.635 | 3.57E-06 |
| DR4 | 7-13 | 1.109 | 0.018 | 5.601 | 2.13E-08 |
| DR4 | 13+ | 1.145 | 0.022 | 6.208 | 5.38E-10 |
| DR3/DR4 | Control | 0.658 | 0.012 | -33.593 | <2.2E-16 |
| DR3/DR4 | 2-7 | 0.910 | 0.014 | -6.865 | 6.63E-12 |

| | | | | | |
|---|---|---|---|---|---|
| DR3/DR4 | 7-13 | 0.870 | 0.013 | -10.386 | <2.2E-16 |
| DR3/DR4 | 13+ | 0.829 | 0.016 | -11.817 | <2.2E-16 |
| DRB1*15:01-DQA1*01:02-DQB1*06:02 | Control | 1.279 | 0.017 | 14.406 | <2.2E-16 |
| DRB1*15:01-DQA1*01:02-DQB1*06:02 | 2-7 | 0.989 | 0.019 | -0.580 | 0.562 |
| DRB1*15:01-DQA1*01:02-DQB1*06:02 | 7-13 | 0.991 | 0.018 | -0.466 | 0.641 |
| DRB1*15:01-DQA1*01:02-DQB1*06:02 | 13+ | 0.996 | 0.022 | -0.176 | 0.861 |
| DRB1*07:01-DQA1*02:01-DQB1*03:03 | Control | 1.086 | 0.010 | 8.442 | <2.2E-16 |
| DRB1*07:01-DQA1*02:01-DQB1*03:03 | 2-7 | 1.009 | 0.011 | 0.838 | 0.402 |
| DRB1*07:01-DQA1*02:01-DQB1*03:03 | 7-13 | 1.011 | 0.011 | 0.999 | 0.318 |
| DRB1*07:01-DQA1*02:01-DQB1*03:03 | 13+ | 1.017 | 0.013 | 1.350 | 0.177 |
| DRB1*13:02-DQA1*01:02-DQB1*06:09 | Control | 1.017 | 0.005 | 3.230 | 0.00124 |
| DRB1*13:02-DQA1*01:02-DQB1*06:09 | 2-7 | 0.998 | 0.006 | -0.300 | 0.765 |
| DRB1*13:02-DQA1*01:02-DQB1*06:09 | 7-13 | 0.998 | 0.006 | -0.354 | 0.723 |

| | | | | | |
|---|---|---|---|---|---|
| DRB1*13:02-DQA1*01:02-DQB1*06:09 | 13+ | 1.004 | 0.007 | 0.645 | 0.519 |
| DRB1*13:01-DQA1*01:03-DQB1*06:03 | Control | 1.081 | 0.011 | 6.872 | 6.31E-12 |
| DRB1*13:01-DQA1*01:03-DQB1*06:03 | 2-7 | 1.007 | 0.013 | 0.583 | 0.56 |
| DRB1*13:01-DQA1*01:03-DQB1*06:03 | 7-13 | 1.012 | 0.012 | 0.954 | 0.34 |
| DRB1*13:01-DQA1*01:03-DQB1*06:03 | 13+ | 1.013 | 0.014 | 0.915 | 0.36 |
| DRB1*08:01-DQA1*04:01-DQB1*04:02 | Control | 0.988 | 0.008 | -1.405 | 0.16 |
| DRB1*08:01-DQA1*04:01-DQB1*04:02 | 2-7 | 1.002 | 0.009 | 0.222 | 0.825 |
| DRB1*08:01-DQA1*04:01-DQB1*04:02 | 7-13 | 0.993 | 0.009 | -0.817 | 0.414 |
| DRB1*08:01-DQA1*04:01-DQB1*04:02 | 13+ | 0.992 | 0.011 | -0.704 | 0.481 |
| HLA A*02:01 | Control | 0.953 | 0.027 | -1.777 | 0.0756 |
| HLA A*02:01 | 2-7 | 1.023 | 0.030 | 0.764 | 0.445 |
| HLA A*02:01 | 7-13 | 1.025 | 0.029 | 0.852 | 0.394 |
| HLA A*02:01 | 13+ | 1.028 | 0.035 | 0.789 | 0.43 |
| HLA A*02:05 | Control | 0.990 | 0.006 | -1.731 | 0.0834 |
| HLA A*02:05 | 2-7 | 1.000 | 0.007 | -0.004 | 0.996 |

| | | | | | |
|---|---|---|---|---|---|
| HLA A*02:05 | 7-13 | 1.000 | 0.007 | -0.019 | 0.985 |
| A*02:05 | 13+ | 0.989 | 0.008 | -1.416 | 0.157 |
| HLA A*11:01 | Control | 1.055 | 0.013 | 4.007 | 6.15E-05 |
| HLA A*11:01 | 2-7 | 1.002 | 0.015 | 0.109 | 0.913 |
| HLA A*11:01 | 7-13 | 1.016 | 0.014 | 1.080 | 0.28 |
| HLA A*11:01 | 13+ | 1.003 | 0.017 | 0.163 | 0.87 |
| HLA B*18:01 | Control | 0.945 | 0.013 | -4.555 | 5.24E-06 |
| HLA B*18:01 | 2-7 | 1.006 | 0.014 | 0.402 | 0.688 |
| HLA B*18:01 | 7-13 | 1.006 | 0.013 | 0.413 | 0.68 |
| HLA B*18:01 | 13+ | 0.983 | 0.016 | -1.082 | 0.279 |
| HLA B*39:06 | Control | 0.911 | 0.007 | -13.374 | <2.2E-16 |
| HLA B*39:06 | 2-7 | 0.974 | 0.008 | -3.334 | 0.000855 |
| HLA B*39:06 | 7-13 | 0.945 | 0.008 | -7.531 | 5.04E-14 |
| HLA B*39:06 | 13+ | 0.928 | 0.009 | -8.306 | <2.2E-16 |
| HLA B*44:03 | Control | 1.078 | 0.013 | 5.826 | 5.68E-09 |
| HLA B*44:03 | 2-7 | 1.011 | 0.014 | 0.732 | 0.464 |
| HLA B*44:03 | 7-13 | 1.009 | 0.014 | 0.652 | 0.514 |
| HLA B*44:03 | 13+ | 1.025 | 0.017 | 1.468 | 0.142 |
| HLA B*45:01 | Control | 1.001 | 0.005 | 0.239 | 0.811 |
| HLA B*45:01 | 2-7 | 0.992 | 0.005 | -1.497 | 0.134 |

| | | | | | |
|---|---|---|---|---|---|
| HLA B*45:01 | 7-13 | 0.996 | 0.005 | -0.823 | 0.411 |
| HLA B*45:01 | 13+ | 1.004 | 0.006 | 0.593 | 0.553 |
| HLA DPB1*03:01 | Control | 0.906 | 0.019 | -5.177 | 2.26E-07 |
| HLA DPB1*03:01 | 2-7 | 1.001 | 0.021 | 0.054 | 0.957 |
| HLA DPB1*03:01 | 7-13 | 0.983 | 0.021 | -0.847 | 0.397 |
| HLA DPB1*03:01 | 13+ | 0.958 | 0.024 | -1.747 | 0.0806 |
| HLA DPB1*04:02 | Control | 1.086 | 0.018 | 4.718 | 2.38E-06 |
| HLA DPB1*04:02 | 2-7 | 0.984 | 0.019 | -0.848 | 0.397 |
| HLA DPB1*04:02 | 7-13 | 0.984 | 0.019 | -0.871 | 0.384 |
| HLA DPB1*04:02 | 13+ | 1.004 | 0.022 | 0.183 | 0.855 |
| HLA DPB1*15:01 | Control | 0.999 | 0.005 | -0.254 | 0.8 |
| HLA DPB1*15:01 | 2-7 | 1.009 | 0.006 | 1.541 | 0.123 |
| HLA DPB1*15:01 | 7-13 | 1.009 | 0.006 | 1.563 | 0.118 |
| HLA DPB1*15:01 | 13+ | 1.007 | 0.007 | 0.978 | 0.328 |
| HLA A*24:02 | Control | 0.903 | 0.017 | -6.122 | 9.25E-10 |
| HLA A*24:02 | 2-7 | 0.993 | 0.018 | -0.381 | 0.703 |
| HLA A*24:02 | 7-13 | 0.957 | 0.018 | -2.469 | 0.0136 |
| HLA A*24:02 | 13+ | 0.883 | 0.021 | -5.844 | 5.1E-09 |
| DR3 | Control | 0.950 | 0.018 | -2.815 | 0.00487 |
| DR3 | 2-7 | 0.970 | 0.020 | -1.548 | 0.122 |

| | | | | | |
|---|---|---|---|---|---|
| DR3 | 7-13 | 0.962 | 0.019 | -2.006 | 0.0448 |
| DR3 | 13+ | 0.967 | 0.023 | -1.474 | 0.14 |
| DR4 | Control | 0.966 | 0.017 | -2.044 | 0.041 |
| DR4 | 2-7 | 1.092 | 0.019 | 4.635 | 3.57E-06 |
| DR4 | 7-13 | 1.109 | 0.018 | 5.601 | 2.13E-08 |
| DR4 | 13+ | 1.145 | 0.022 | 6.208 | 5.38E-10 |
| DR3/DR4 | Control | 0.658 | 0.012 | -33.593 | <2.2E-16 |
| DR3/DR4 | 2-7 | 0.910 | 0.014 | -6.865 | 6.63E-12 |
| DR3/DR4 | 7-13 | 0.870 | 0.013 | -10.386 | <2.2E-16 |
| DR3/DR4 | 13+ | 0.829 | 0.016 | -11.817 | <2.2E-16 |
| DRB1*15:01-DQA1*01:02-DQB1*06:02 | Control | 1.279 | 0.017 | 14.406 | <2.2E-16 |
| DRB1*15:01-DQA1*01:02-DQB1*06:02 | 2-7 | 0.989 | 0.019 | -0.580 | 0.562 |
| DRB1*15:01-DQA1*01:02-DQB1*06:02 | 7-13 | 0.991 | 0.018 | -0.466 | 0.641 |
| DRB1*15:01-DQA1*01:02-DQB1*06:02 | 13+ | 0.996 | 0.022 | -0.176 | 0.861 |
| DRB1*07:01-DQA1*02:01-DQB1*03:03 | Control | 1.086 | 0.010 | 8.442 | <2.2E-16 |
| DRB1*07:01-DQA1*02:01-DQB1*03:03 | 2-7 | 1.009 | 0.011 | 0.838 | 0.402 |

| | | | | | |
|---|---|---|---|---|---|
| DRB1*07:01-DQA1*02:01-DQB1*03:03 | 7-13 | 1.011 | 0.011 | 0.999 | 0.318 |
| DRB1*07:01-DQA1*02:01-DQB1*03:03 | 13+ | 1.017 | 0.013 | 1.350 | 0.177 |
| DRB1*13:02-DQA1*01:02-DQB1*06:09 | Control | 1.017 | 0.005 | 3.230 | 0.00124 |
| DRB1*13:02-DQA1*01:02-DQB1*06:09 | 2-7 | 0.998 | 0.006 | -0.300 | 0.765 |
| DRB1*13:02-DQA1*01:02-DQB1*06:09 | 7-13 | 0.998 | 0.006 | -0.354 | 0.723 |
| DRB1*13:02-DQA1*01:02-DQB1*06:09 | 13+ | 1.004 | 0.007 | 0.645 | 0.519 |
| DRB1*13:01-DQA1*01:03-DQB1*06:03 | Control | 1.081 | 0.011 | 6.872 | 6.31E-12 |
| DRB1*13:01-DQA1*01:03-DQB1*06:03 | 2-7 | 1.007 | 0.013 | 0.583 | 0.56 |
| DRB1*13:01-DQA1*01:03-DQB1*06:03 | 7-13 | 1.012 | 0.012 | 0.954 | 0.34 |
| DRB1*13:01-DQA1*01:03-DQB1*06:03 | 13+ | 1.013 | 0.014 | 0.915 | 0.36 |
| DRB1*08:01-DQA1*04:01-DQB1*04:02 | Control | 0.988 | 0.008 | -1.405 | 0.16 |
| DRB1*08:01-DQA1*04:01-DQB1*04:02 | 2-7 | 1.002 | 0.009 | 0.222 | 0.825 |

| | | | | | |
|---|---|---|---|---|---|
| DRB1*08:01-DQA1*04:01-DQB1*04:02 | 7-13 | 0.993 | 0.009 | -0.817 | 0.414 |
| DRB1*08:01-DQA1*04:01-DQB1*04:02 | 13+ | 0.992 | 0.011 | -0.704 | 0.481 |
| HLA A*02:01 | Control | 0.953 | 0.027 | -1.777 | 0.0756 |
| HLA A*02:01 | 2-7 | 1.023 | 0.030 | 0.764 | 0.445 |
| HLA A*02:01 | 7-13 | 1.025 | 0.029 | 0.852 | 0.394 |
| HLA A*02:01 | 13+ | 1.028 | 0.035 | 0.789 | 0.43 |
| HLA A*02:05 | Control | 0.990 | 0.006 | -1.731 | 0.0834 |
| HLA A*02:05 | 2-7 | 1.000 | 0.007 | -0.004 | 0.996 |
| HLA A*02:05 | 7-13 | 1.000 | 0.007 | -0.019 | 0.985 |
| HLA A*02:05 | 13+ | 0.989 | 0.008 | -1.416 | 0.157 |
| HLA A*11:01 | Control | 1.055 | 0.013 | 4.007 | 6.15E-05 |
| HLA A*11:01 | 2-7 | 1.002 | 0.015 | 0.109 | 0.913 |
| HLA A*11:01 | 7-13 | 1.016 | 0.014 | 1.080 | 0.28 |
| HLA A*11:01 | 13+ | 1.003 | 0.017 | 0.163 | 0.87 |
| HLA B*18:01 | Control | 0.945 | 0.013 | -4.555 | 5.24E-06 |
| HLA B*18:01 | 2-7 | 1.006 | 0.014 | 0.402 | 0.688 |
| HLA B*18:01 | 7-13 | 1.006 | 0.013 | 0.413 | 0.68 |
| HLA B*18:01 | 13+ | 0.983 | 0.016 | -1.082 | 0.279 |
| HLA B*39:06 | Control | 0.911 | 0.007 | -13.374 | <2.2E-16 |

| | | | | | |
|---|---|---|---|---|---|
| HLA B*39:06 | 2-7 | 0.974 | 0.008 | -3.334 | 0.000855 |
| HLA B*39:06 | 7-13 | 0.945 | 0.008 | -7.531 | 5.04E-14 |
| HLA B*39:06 | 13+ | 0.928 | 0.009 | -8.306 | <2.2E-16 |
| HLA B*44:03 | Control | 1.078 | 0.013 | 5.826 | 5.68E-09 |
| HLA B*44:03 | 2-7 | 1.011 | 0.014 | 0.732 | 0.464 |
| HLA B*44:03 | 7-13 | 1.009 | 0.014 | 0.652 | 0.514 |
| HLA B*44:03 | 13+ | 1.025 | 0.017 | 1.468 | 0.142 |
| HLA B*45:01 | Control | 1.001 | 0.005 | 0.239 | 0.811 |
| HLA B*45:01 | 2-7 | 0.992 | 0.005 | -1.497 | 0.134 |
| HLA B*45:01 | 7-13 | 0.996 | 0.005 | -0.823 | 0.411 |
| HLA B*45:01 | 13+ | 1.004 | 0.006 | 0.593 | 0.553 |
| HLA DPB1*03:01 | Control | 0.906 | 0.019 | -5.177 | 2.26E-07 |
| HLA DPB1*03:01 | 2-7 | 1.001 | 0.021 | 0.054 | 0.957 |
| HLA DPB1*03:01 | 7-13 | 0.983 | 0.021 | -0.847 | 0.397 |
| HLA DPB1*03:01 | 13+ | 0.958 | 0.024 | -1.747 | 0.0806 |
| HLA DPB1*04:02 | Control | 1.086 | 0.018 | 4.718 | 2.38E-06 |
| HLA DPB1*04:02 | 2-7 | 0.984 | 0.019 | -0.848 | 0.397 |
| HLA DPB1*04:02 | 7-13 | 0.984 | 0.019 | -0.871 | 0.384 |
| HLA DPB1*04:02 | 13+ | 1.004 | 0.022 | 0.183 | 0.855 |
| HLA DPB1*15:01 | Control | 0.999 | 0.005 | -0.254 | 0.8 |

| | | | | | |
|---|---|---|---|---|---|
| HLA DPB1*15:01 | 2-7 | 1.009 | 0.006 | 1.541 | 0.123 |
| HLA DPB1*15:01 | 7-13 | 1.009 | 0.006 | 1.563 | 0.118 |
| HLA DPB1*15:01 | 13+ | 1.007 | 0.007 | 0.978 | 0.328 |
