## Supplementary Table 5 for "Genetic risk in extremely early onset type 1 diabetes"

**Supplementary Table 5.** Regression output comparing HLA contribution to Control group including interaction terms.

| Predictor | <2 OR | <2 SE | <2 *P*-value | 2-7 OR | 2-7 SE | 2-7 *P*-value | 7-13 OR | 7-13 SE | 7-13 *P*-value | 13+ OR | 13+ SE | 13+ *P*-value |
|---|---|---|---|---|---|---|---|---|---|---|---|---|
| A*24:02 | 1.06 | 0.39 | 8.85E-01 | 2.27 | 0.14 | 2.73E-09 | 1.41 | 0.12 | 3.45E-03 | 0.81 | 0.26 | 4.20E-01 |
| A*24:02 × B*39:06 | 1.62 | 0.33 | 1.39E-01 | 0.97 | 0.22 | 8.73E-01 | 0.98 | 0.22 | 9.12E-01 | 0.95 | 0.40 | 9.00E-01 |
| A*24:02 × DR3-DQ2/DR3-DQ2 | 1.84 | 0.52 | 2.43E-01 | 1.32 | 0.35 | 4.24E-01 | 1.75 | 0.34 | 1.00E-01 | 0.26 | 1.08 | 2.16E-01 |
| A*24:02 × DR3-DQ2/DR4-DQ8 | 1.25 | 0.44 | 6.03E-01 | 0.66 | 0.22 | 6.54E-02 | 0.85 | 0.21 | 4.53E-01 | 0.84 | 0.38 | 6.43E-01 |
| A*24:02 × DR3-DQ2/X | 1.90 | 0.45 | 1.57E-01 | 1.09 | 0.19 | 6.59E-01 | 1.74 | 0.17 | 1.12E-03 | 1.49 | 0.36 | 2.66E-01 |
| A*24:02 × DR4-DQ8/DR4-DQ8 | 1.05 | 0.77 | 9.53E-01 | 0.64 | 0.34 | 1.88E-01 | 0.92 | 0.33 | 7.88E-01 | 0.79 | 0.53 | 6.51E-01 |
| A*24:02 × DR4-DQ8/X | 1.72 | 0.44 | 2.18E-01 | 0.46 | 0.18 | 2.58E-05 | 0.80 | 0.16 | 1.61E-01 | 1.12 | 0.31 | 7.15E-01 |

| | | | | | | | | | | | | |
|---|---|---|---|---|---|---|---|---|---|---|---|---|
| B*39:06 | 9.83 | 0.51 | 6.97E-06 | 12.31 | 0.24 | <2.2E-16 | 6.92 | 0.23 | <2.2E-16 | 7.14 | 0.37 | 1.59E-07 |
| B*39:06 × DR3-DQ2/DR3-DQ2 | 1.06e+59 | 0.59 | <2.2E-16 | 4.85e+57 | 0.73 | <2.2E-16 | 6.16e+58 | 0.48 | <2.2E-16 | 9.61E-27 | 9.16E-86 | <2.2E-16 |
| B*39:06 × DR3-DQ2/DR4-DQ8 | 0.17 | 0.65 | 6.51E-03 | 0.12 | 0.49 | 1.22E-05 | 0.17 | 0.49 | 4.22E-04 | 0.05 | 1.14 | 9.03E-03 |
| B*39:06 × DR3-DQ2/X | 0.72 | 0.59 | 5.75E-01 | 0.52 | 0.31 | 4.05E-02 | 0.48 | 0.32 | 2.21E-02 | 0.25 | 0.64 | 3.13E-02 |
| B*39:06 × DR4-DQ8/DR4-DQ8 | 3.82E-59 | NA | NA | 0.06 | 0.69 | 2.91E-05 | 0.04 | 0.79 | 7.04E-05 | 0.08 | 1.12 | 2.64E-02 |
| B*39:06 × DR4-DQ8/X | 0.81 | 0.55 | 7.07E-01 | 0.46 | 0.30 | 1.04E-02 | 0.41 | 0.30 | 3.64E-03 | 0.37 | 0.50 | 4.62E-02 |
| DR3-DQ2/DR3-DQ2 | 64.21 | 0.24 | <2.2E-16 | 33.88 | 0.14 | <2.2E-16 | 18.57 | 0.12 | <2.2E-16 | 18.71 | 0.17 | <2.2E-16 |
| DR3-DQ2/DR4-DQ8 | 141.56 | 0.21 | <2.2E-16 | 88.77 | 0.11 | <2.2E-16 | 46.37 | 0.09 | <2.2E-16 | 35.04 | 0.14 | <2.2E-16 |
| DR3-DQ2/X | 6.64 | 0.23 | <2.2E-16 | 5.22 | 0.11 | <2.2E-16 | 3.07 | 0.08 | <2.2E-16 | 3.07 | 0.14 | 2.22E-16 |
| DR4-DQ8/DR4-DQ8 | 31.95 | 0.36 | <2.2E-16 | 49.41 | 0.17 | <2.2E-16 | 26.93 | 0.15 | <2.2E-16 | 30.17 | 0.20 | <2.2E-16 |

| | | | | | | | | | | | | |
|---|---|---|---|---|---|---|---|---|---|---|---|---|
| DR4-DQ8/X | 7.67 | 0.23 | <2.2E-16 | 10.84 | 0.11 | <2.2E-16 | 7.06 | 0.08 | <2.2E-16 | 6.65 | 0.13 | <2.2E-16 |
