## Supplementary Table 6 for "Genetic risk in extremely early onset type 1 diabetes"

**Supplementary Table 6.** Discriminative Performance of T1D-GRS Across Population Centile Thresholds type 1 diabetes diagnosed <2 years in EXE-T1D/EXTEND/PRB cohort and T1DGC cohort combined. Risk of type 1 diabetes diagnosed <2 years calculated using a 0.00012% population prevalence (26). Population centile calculated from UK Biobank European population.

| Population Centile | T1D Centile | T1D-GRS | Sensitivity (%) | Specificity (%) | 1-Specificity (%) | Youden index | T1D risk (%) |
|---|---|---|---|---|---|---|---|
| 50 | 1.9 | 10.159 | 98.1 | 44.1 | 55.9 | 0.422 | 0.02 |
| 75 | 3.4 | 11.815 | 96.6 | 69.0 | 31.0 | 0.657 | 0.04 |
| 80 | 3.7 | 12.185 | 96.3 | 74.3 | 25.7 | 0.706 | 0.04 |
| 85 | 5.7 | 12.599 | 94.3 | 79.9 | 20.1 | 0.742 | 0.06 |
| 90 | 8.9 | 13.115 | 91.1 | 86.0 | 14.0 | 0.771 | 0.08 |
| 95 | 20.5 | 13.882 | 79.5 | 92.3 | 7.7 | 0.718 | 0.12 |
| 99 | 51 | 15.303 | 49.0 | 98.0 | 2.0 | 0.470 | 0.29 |
| 100 | 100 | 19.103 | 0 | 100 | 0 | 0 | NA |
