## Supplementary Table 7 for "Genetic risk in extremely early onset type 1 diabetes"

**Supplementary Table 7.** Discriminative Performance of T1D-GRS across Population Centile Thresholds for type 1 diabetes diagnosed (T1D) <2 years in A) EXE-T1D/EXTEND/PRB cohort. Population centile calculated from UK Biobank European population.

| Population Centile | T1D Centile | T1D-GRS | Sensitivity (%) | Specificity (%) | 1-Specificity (%) | Youden index |
|---|---|---|---|---|---|---|
| 50 | 3.7 | 10.159 | 96.3 | 42.9 | 57.1 | 0.392 |
| 75 | 5.8 | 11.815 | 94.2 | 67.6 | 32.4 | 0.618 |
| 80 | 6.2 | 12.185 | 93.8 | 72.8 | 27.2 | 0.666 |
| 85 | 9.1 | 12.599 | 90.9 | 78.8 | 21.2 | 0.697 |
| 90 | 12.0 | 13.115 | 88.0 | 85.1 | 14.9 | 0.731 |
| 95 | 25.6 | 13.882 | 74.4 | 91.7 | 8.3 | 0.661 |
| 99 | 55.8 | 15.303 | 44.2 | 98.0 | 2.0 | 0.422 |
| 100 | 100 | 19.103 | 0 | 100 | 0 | 0 |
