## Supplementary Table 8 for "Genetic risk in extremely early onset type 1 diabetes"

**Supplementary Table 8.** Discriminative Performance of T1D-GRS across Population Centile Thresholds for type 1 diabetes diagnosed (T1D) <2 years in T1DGC cohort. Population centile calculated from UK Biobank European population.

| Population Centile | T1D Centile | T1D-GRS | Specificity (%) | Sensitivity (%) | 1-Specificity (%) | Youden index |
|---|---|---|---|---|---|---|
| 50 | 0.6 | 10.159 | 99.4 | 45.2 | 54.8 | 0.447 |
| 75 | 1.7 | 11.815 | 98.3 | 70.6 | 29.4 | 0.689 |
| 80 | 2.0 | 12.185 | 98.0 | 75.9 | 24.1 | 0.739 |
| 85 | 3.4 | 12.599 | 96.6 | 81.2 | 18.8 | 0.778 |
| 90 | 6.8 | 13.115 | 93.2 | 87.0 | 13.0 | 0.802 |
| 95 | 17.0 | 13.882 | 83.0 | 93.0 | 7.0 | 0.759 |
| 99 | 47.7 | 15.303 | 52.3 | 98.0 | 2.0 | 0.503 |
| 100 | 100 | 19.103 | 0 | 100 | 0 | 0 |
