## Supplementary Fig. 1 for "Genetic risk in extremely early onset type 1 diabetes"

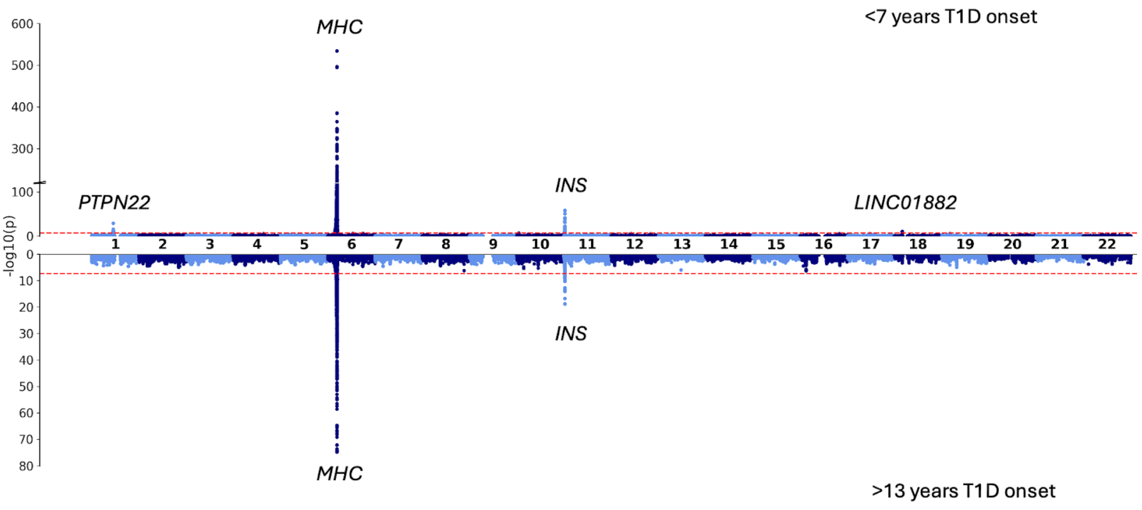


**Supplementary Figure 1.** Miami plot for variants associated with <7 years type 1 diabetes (T1D) onset (top) and >13 years T1D onset (bottom). Red line represents *P*=5x10$^{-8}$. Loci are labelled based on nearest gene.
