## Supplementary Fig. 2 for "Genetic risk in extremely early onset type 1 diabetes"

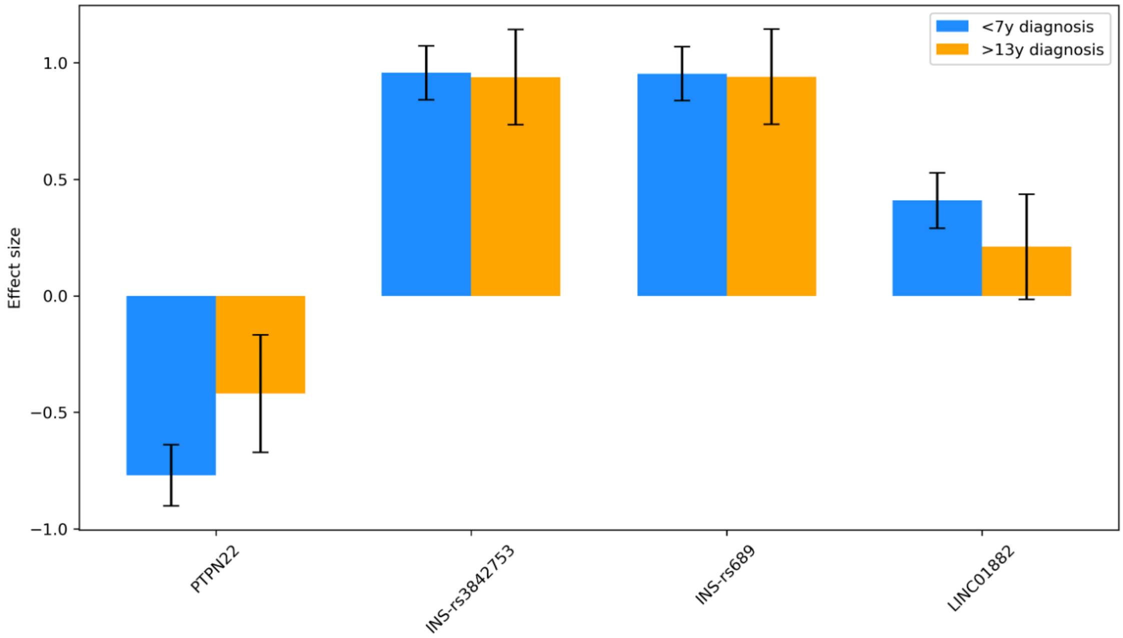


**Supplementary Figure 2.** Effect sizes of variants significant in association testing for <7 years (blue) and >13 years (orange) type 1 diabetes onset. Error bars represent 95% confidence intervals.
