## Supplementary Fig. 3 for "Genetic risk in extremely early onset type 1 diabetes"

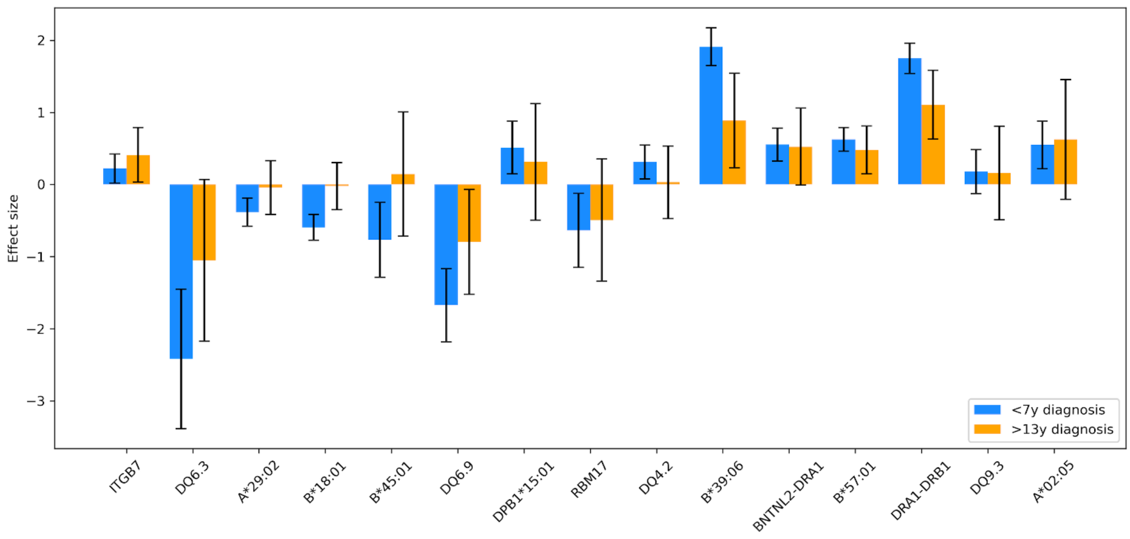


**Supplementary Figure 3.** Effect sizes for nominally significant (P<0.05) T1D-GRS for <7 years (blue) and >13 years (orange) type 1 diabetes onset. *= P<0.05. Error bars represent 95% confidence intervals.
