## Supplementary Fig. 4 for "Genetic risk in extremely early onset type 1 diabetes"

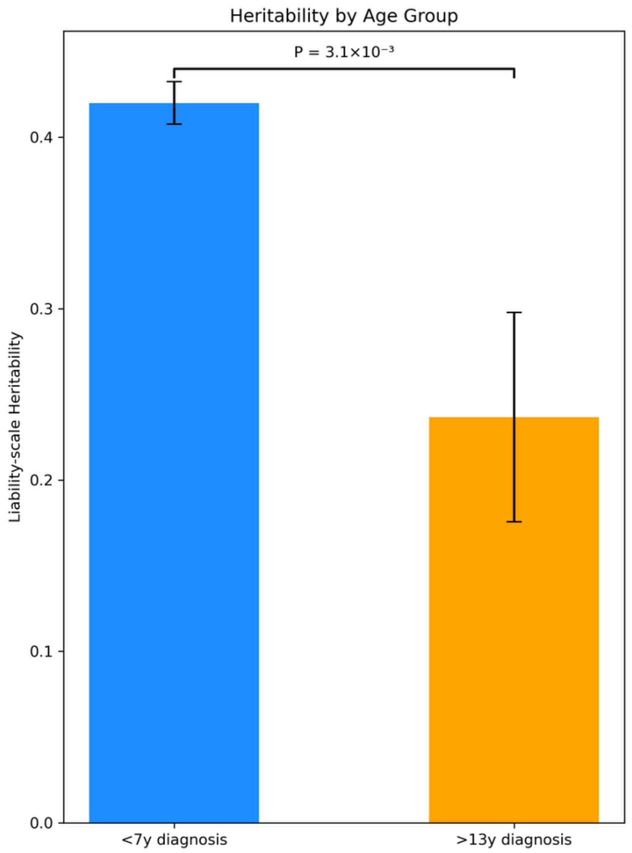


**Supplementary Figure 4.** Liability-scale heritability of <7 years onset (blue) and >13 years onset (orange). Error bars represent standard error.
