## Supplementary Fig. 5 for "Genetic risk in extremely early onset type 1 diabetes"

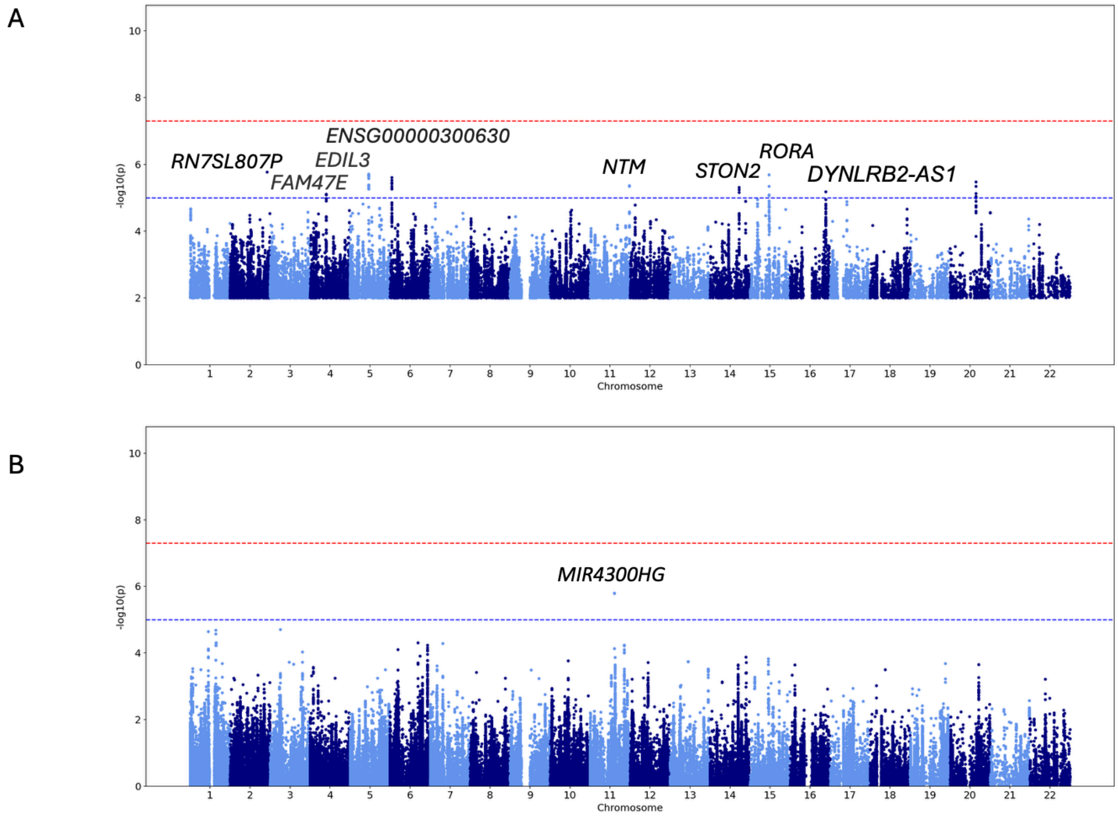


**Supplementary Figure 5.** Manhattan plots for continuous phenotype age of diagnosis for A) EXET1D/EXTEND/PRB cohorts and B) T1DGC cohort. Red line represents $P$=5x10$^{-8}$, blue line represents p=1x10$^{-5}$. Loci are labelled based on nearest gene.
