## Supplementary Fig. 6 for "Genetic risk in extremely early onset type 1 diabetes"

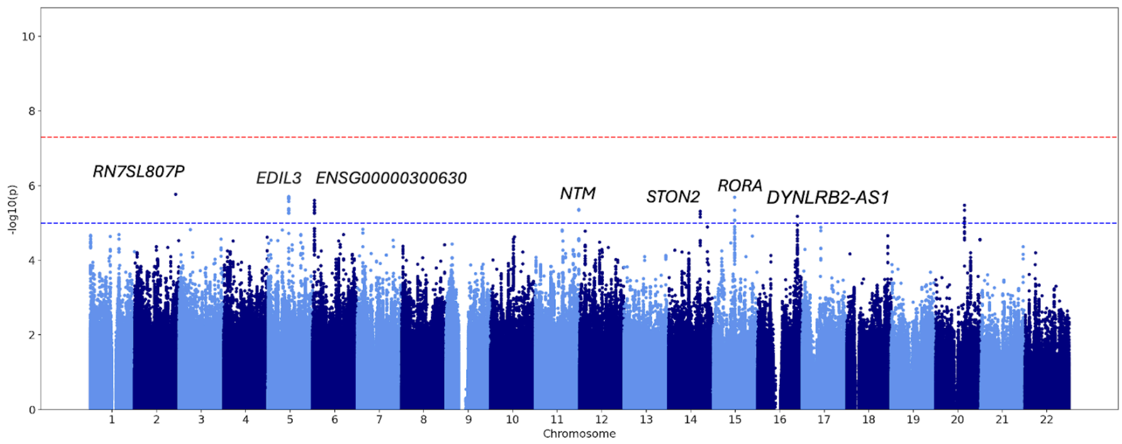


**Supplementary Figure 6.** Manhattan plots from meta-analysis of continuous phenotype age of diagnosis for EXET1D/EXTEND/PRB cohorts and T1DGC cohort. Red line represents $P$=5x10$^{-8}$, blue line represents p=1x10$^{-5}$. Loci are labelled based on nearest gene.
