## Supplementary Fig. 7 for "Genetic risk in extremely early onset type 1 diabetes"

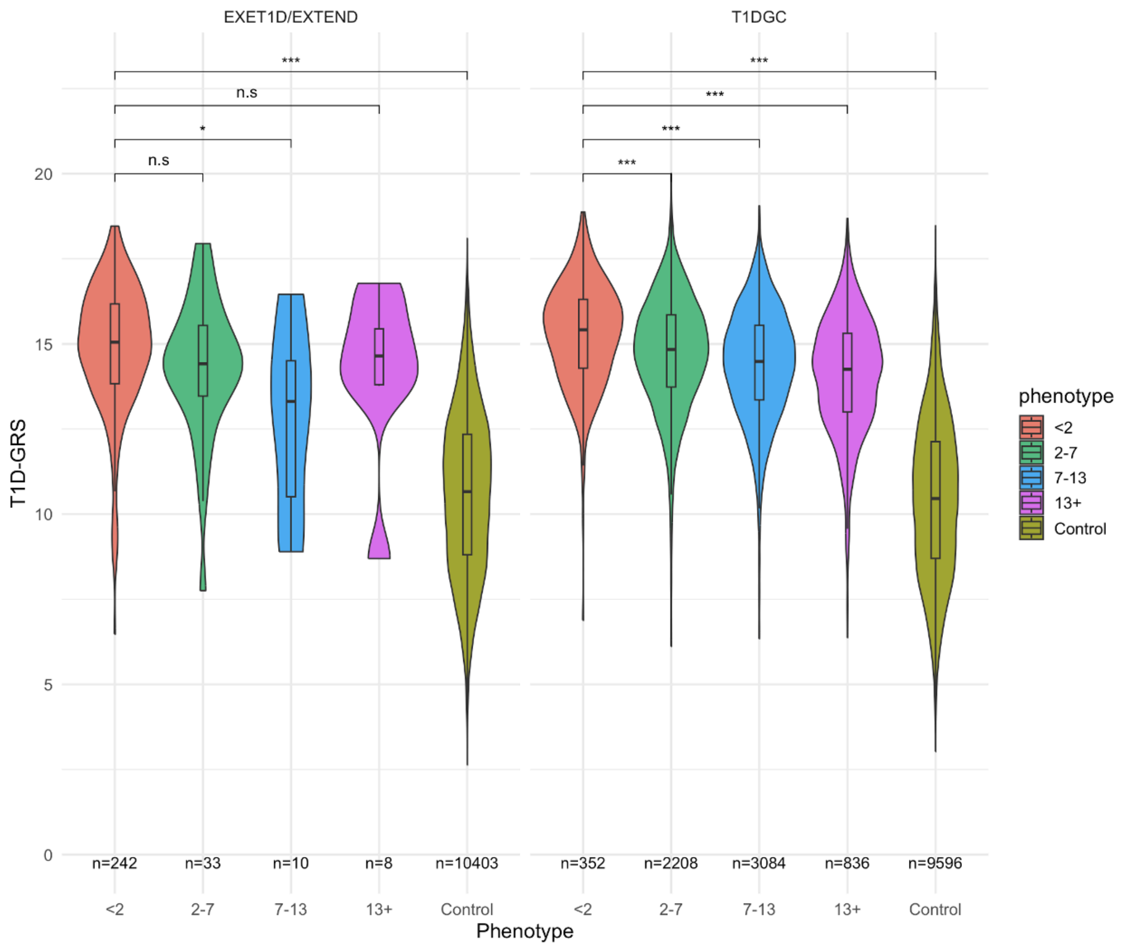


**Supplementary Figure 7.** T1D-GRS Distribution across different diagnosis groupings in T1DGC and EXE-T1D/EXTEND/PRB Cohorts. Violin plots show the distribution of the 67-SNP T1D-GRS for type 1 diabetes cases stratified by age of diagnosis compared to controls in both the EXE-T1D/EXTEND/PRB (left) and T1DGC (right) cohorts. In the T1DGC cohort, individuals diagnosed before 2 years of age had the highest T1D-GRS (mean=15.3 [95% CI 15.1–15.4]) compared with those diagnosed at 2–7 years (mean=14.7 [95% CI 14.7–14.8]; *Padj*=7.2 x$10^{-6}$), 7–13 years (mean=14.4 [95% CI 14.3–14.5]; *Padj*= 1.39x$10^{-13}$), and >13 years (mean=14.1 [95% CI 14.0–14.2]; *Padj*=1.2x$10^{-15}$) (Fig. 2). In the EXE-T1D/EXTEND/PRB cohort, individuals with onset <2 years had a mean T1D-GRS of 14.9 (95% CI 14.7–15.1) (Fig. 2). This did not differ significantly from onset at 2–7 years (mean=14.3 [95% CI 13.6–15.0]; *Padj*=0.28) or >13 years (mean=14.2 [95% CI 12.1–16.3]; *Padj*=0.31, but was significantly higher than in those with onset at 7–13 years (mean=12.8 [95% CI 10.8–14.8]; *Padj*=1.7x$10^{-2}$). In both cohorts, the control group had the lowest T1D-GRS (EXE-T1D/EXTEND/PRB mean=10.6 [95% CI 10.5–10.6]; T1DGC mean=10.5 [95% CI 10.4–10.5]; *Padj*=1.2

$x10^{-15}$ for both). The box plot within each violin shows the median, interquartile range (IQR), and 95% confidence intervals. All *P*-values derived from logistic regression adjusting for first five Principal Components (PCs) and sex. *** = $P<0.001$, *= $P<0.05$, n.s = $P>0.05$
