## Supplementary Fig. 8 for "Genetic risk in extremely early onset type 1 diabetes"

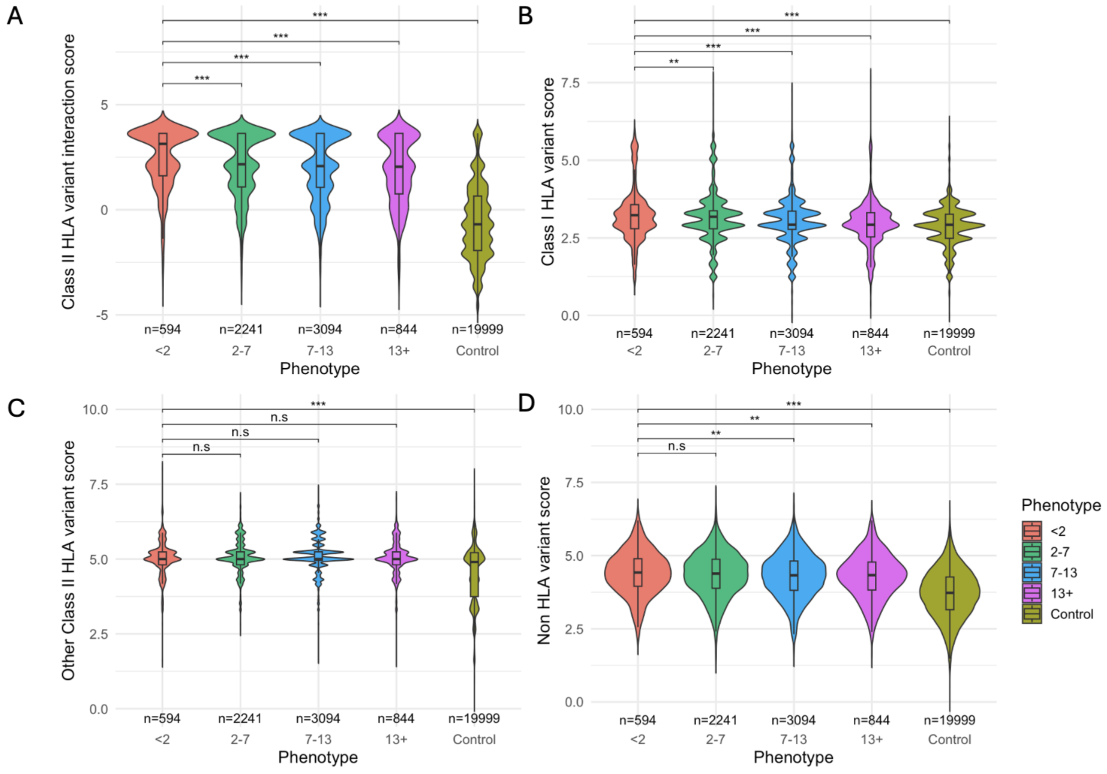


**Supplementary Figure 8.** Partitioned polygenic T1D-GRS distribution across different diagnosis groupings in T1DGC and EXE-T1D/EXTEND/PRB Cohorts. A) Individuals diagnosed <2 years had a higher mean Class II HLA interaction score (2.46 95% CI 2.36-2.57) than those diagnosed: 2-7 years (2.14 95% CI 2.08-2.21; *Padj*=5.63x$10^{-6}$); 7-13 years (1.97 95% CI 1.91-2.02; *Padj*=4.84x$10^{-12}$); 13+ years (1.82 95% CI 1.71-1.93; *Padj*=1.92x$10^{-13}$); and compared to the control group (-0.56 95% CI -0.59- -0.54 ; *Padj*= 8.0x$10^{-16}$). B) Individuals diagnosed <2 years had a higher mean Class I HLA interaction score (3.23 95% CI 3.17-3.30) than those diagnosed: 2-7 years (3.14 95% CI 3.11-3.18; *Padj*=1.94x$10^{-3}$); 7-13 years (3.04 95% CI 3.01-3.07; *Padj*=7.92x$10^{-11}$); 13+ years (2.95 95% CI 2.90-3.00; *Padj*=2.28x$10^{-15}$); and compared to the control group (2.87 95% CI 2.86- 2.88; *Padj*= 8.0x$10^{-16}$). C) We found no significant difference (*P*>0.05) between different ages of onset when comparing Class 2 HLA variants (excluding interaction terms) (<2 years onset score= 5.04, 95% CI 4.99-5.08; 2-7 years onset score= 5.07, 95% CI 5.05-5.09; 7-13 years onset score= 5.09, 95% CI 5.07-5.11; 13 years+ score= 5.04, 95% CI= 5.00-5.08); though all age groups had a higher score than the control group (Control group score= 4.52, 95% CI 4.51-4.53; *Padj*= 8.0x$10^{-16}$). D) Individuals diagnosed <2 years had a higher mean non-HLA (4.40 95% CI 4.34-4.46) variant score than those diagnosed: 7-13 years (4.35, 95% CI 4.27-4.33; *Padj*= $3.94 \times 10^{-3}$); 13 years+ (4.29, 95% CI 4.24-4.34; *Padj*= $4.41 \times 10^{-3}$); and the control group (3.70, 95% CI 3.69-3.71; *Padj*= $8 \times 10^{-16}$). Whilst <2 years also had a higher score than those diagnosed 2-7 years (4.35, 95% CI 4.32-4.39) this was not statistically significant (*Padj*= $1.01 \times 10^{-1}$). The box plot within each violin shows the median, interquartile range (IQR), and 95% confidence intervals. All *P*-values derived from logistic regression adjusting for first five Principal Components (PCs), sex, and array used. *** = *P*<0.001, *= *P*<0.05, n.s = P>0.05.
