## Supplementary Fig. 9 for "Genetic risk in extremely early onset type 1 diabetes"

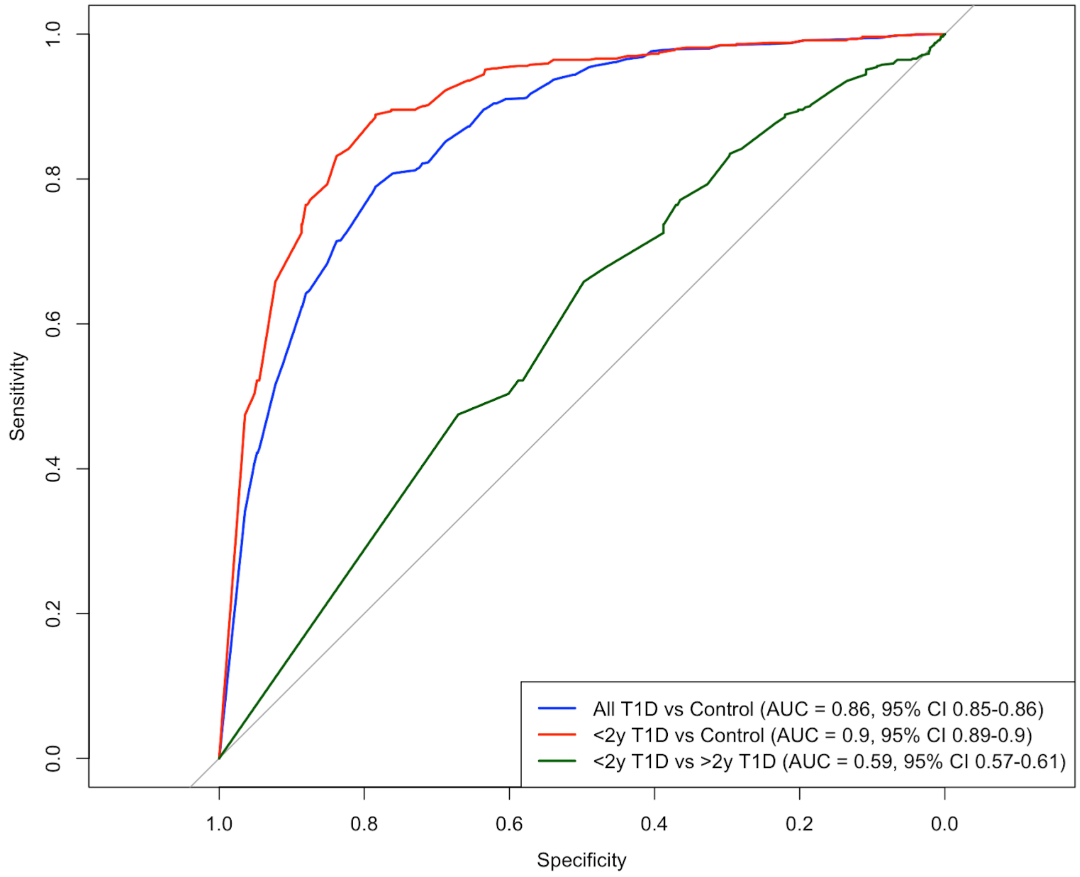


**Supplementary Figure 9.** Receiver Operating Characteristic (ROC) Curves for partitioned Class II HLA variant score. ROC curves of the Class II HLA variant score discriminative ability between type 1 diabetes (T1D) phenotypes: all T1D cases vs. controls (AUC=0.86, 95% CI 0.85-0.86) (blue), extremely early-onset T1D (<2 years) vs. controls (red) (AUC=0.9, 95% CI 0.89-0.9), <2 years T1D vs. >2 years T1D onset (green) (AUC=0.59, 95% CI 0.57-0.61).
